# The genetic architecture of wisdom and its relationship with health, personality and cognition

**DOI:** 10.64898/2026.11.10.26367758

**Authors:** Fabrizio Malatesta, Hayley Thorpe, Jean Gonzalez, John J Meredith, Laura Vilar-Ribó, Natasia Courchesne-Krak, Daniel Gustavson, 23andMe Research Team, Vinh Tran, Shubham Saini, Emily M. Rios, Abraham Palmer, Sandra Sanchez-Roige

**Affiliations:** Department of Psychiatry, University of California San Diego, La Jolla, CA, USA; Department of Anatomy and Cell Biology, Schulich School of Medicine and Dentistry, Western University, London, ON, Canada; Institute for Behavioral Genetics, University of Colorado Boulder, CO, USA; Department of Psychology and Neuroscience, University of Colorado Boulder, CO, USA; 23andMe Research Institute, Palo Alto, CA, USA; Institute for Genomic Medicine, University of California San Diego, La Jolla, CA, USA; Department of Medicine, Vanderbilt University Medical Center, Nashville, TN, USA

## Abstract

Wisdom is a personality trait composed of self-reflection, pro-social behaviors, emotional regulation, acceptance of divergent perspectives, decisiveness, social advising, and spirituality. Greater levels of wisdom are linked to better overall health, well-being, happiness, life satisfaction, and resilience. However, the underlying genetic architecture of wisdom remains unexplored. We assessed wisdom using responses to the SD-WISE-7 in 153,384 23andMe research-consented participants and conducted a multi-ancestry GWAS. We identified one genome-wide significant locus (rs2325631, *p*=1.41E-08) near *MANEA* on chromosome 6. Wisdom was heritable (*h^2^_SNP_*=0.03±4.50E-03) and genetically correlated with numerous traits, including greater subjective well-being, openness, and risk tolerance, and lower neuroticism and loneliness. Crucially, wisdom was genetically distinct from intelligence and the cognitive component of educational attainment. Wisdom polygenic scores were associated with psychosocial outcomes, but not cognitive performance in All of Us. This study advances our understanding of wisdom as a heritable trait that could guide interventions to improve human health.

## Introduction

Wisdom is a personality trait, which has been broadly defined as a multidimensional integration of cognitive insight, self-reflection, and prosocial behavior (such as empathy and compassion)^1^, and is a correlate of health and well-being^2^. Wisdom is distinct from intelligence^3,4^, which broadly involves reasoning, abstraction, problem-solving, and learning from experience^5^. Among the most widely used instruments for assessing wisdom are the 24-item San Diego Wisdom Scale (**SD-WISE-24**^6^) and the shorter 7-item version derived from the expanded scale (**SD-WISE-7**^7^), which captures 1) Self-Reflection, 2) Pro-Social Behaviors, 3) Emotional Regulation, 4) Acceptance of Divergent Perspectives, 5) Decisiveness, 6) Social Advising, and 7) Spirituality. Prior studies have shown that greater levels of wisdom are positively associated with mental and physical health^1,8,9^, happiness^10^, life satisfaction^11^, and resilience^9^, and negatively associated with loneliness^12^, depression, and anxiety^9^. Components of wisdom also have potential relevance to behavioral symptoms of neuropsychiatric disorders that have disrupted judgment and personality^13^, such as frontotemporal dementia^,14^ and autism spectrum disorder^15^. Longitudinal studies show that wisdom increases with age^16,17^ and is potentially modifiable, with behavioral intervention studies reporting increases in wisdom or specific wisdom components^18–20^. This suggests wisdom’s potential application to interventions that improve health outcomes.

The biological underpinnings of wisdom have been previously theorized^2,13^. The evolutionary Grandmother Hypothesis proposes that post-reproductive adults benefit their kin by investing effort into grandchildren, allowing their children to reproduce sooner^21^. Several authors have extended this idea to wisdom, proposing that knowledge and wisdom accumulated through experience likewise allow older adults to benefit their kin^2,22,23^. However, the genetic underpinnings of wisdom have not been studied, despite other related traits being heritable, such as subjective well-being^24^, resilience^25^, loneliness^26^, and Big Five personality traits Big Five personality traits (i.e., extraversion, agreeableness, conscientiousness, neuroticism, openness to experience)^27^. Elucidating the genetic underpinnings of wisdom can therefore help our understanding of the biological basis of wisdom, enhance diagnostic mechanisms and develop biologically-informed interventions to increase wisdom.

Genome-wide association studies (**GWAS**) offer a powerful way to identify the genetic basis of wisdom and its relationships with health, as has been done for other personality and behavioral traits^26–29^. We collected SD-WISE-7 data from a large cohort of genotyped research-consented participants (*N* = 161,775) from the 23andMe Research Institute. Our objectives were four-fold. First, we examined phenotypic correlations between wisdom and demographic, socioeconomic status (**SES**), behavioral, psychiatric, and physical health measures. Second, we performed the first GWAS of wisdom, and calculated its heritability. Third, using Linkage Disequilibrium Score Regression (**LDSC**), we estimated genetic correlations between wisdom and a broad range of demographic/socioeconomic, behavioral and personality, externalizing and risk-behavior, psychiatric, physical health, neuroimaging, and cognitive traits from publicly available GWAS. We also estimated genetic correlations with intelligence, which is conceptually related to but distinct from wisdom^3^. Fourth, we calculated polygenic scores (**PGS**) in the All of Us Research Program (**AoU**) to test the association between genetic liability of wisdom and measures of mental health, psychosocial and physical outcomes, well-being, as well as task-based cognitive performance. By integrating psychometric data with large-scale genomic analysis, this study advances our understanding of wisdom as a biologically grounded trait and clarifies its genetic relationship to personality, psychopathology, well-being, and cognition.

## Results

### Participant Demographics and SD-WISE-7 Measures

As part of a larger study questionnaire on health and lifestyle^30,31^, we collected SD-WISE-7 responses for 161,775 research-consented participants across the US. Participant demographics and SD-WISE-7 score distributions are described in **Figure 1** and **Supplementary Tables 1-2**. The cohort was 70.7% female and a mean age of 52.9 ± 16.0 (range 18-90) years old.

**Figure 1.**
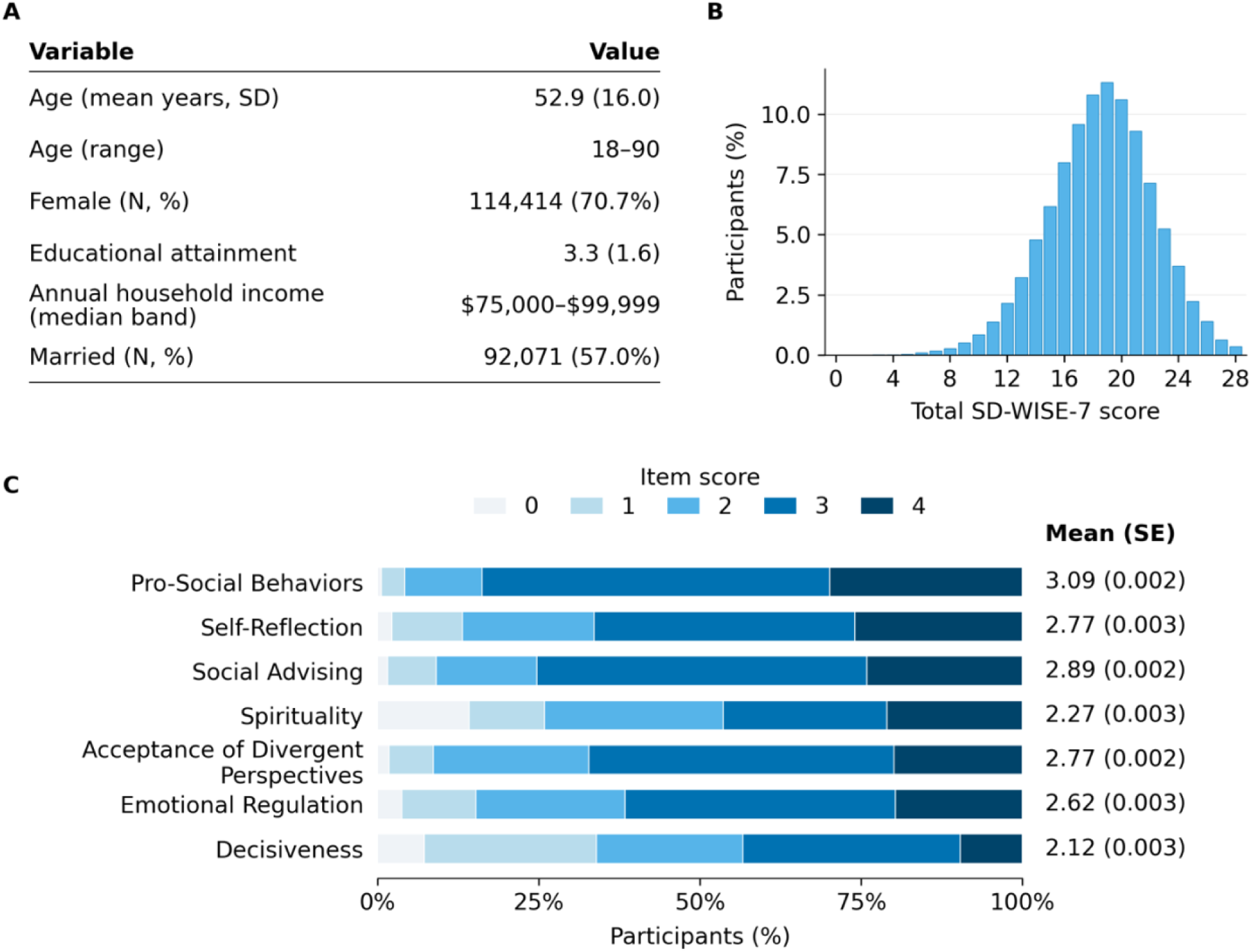
Participant demographics and SD-WISE-7 score distributions. **A)** Table showing demographic variables, including age, sex at birth (male, female), educational attainment, household income, marital status. Educational attainment categories were: less than high school (0), high school diploma (1), some college (2), associates degree (3), bachelors degree (4), masters degree (5), professional degree (6), doctorate degree (7). Household income categories were: <$10,000, $10,000-24,999, $25,000-34,999, $35,000-49,999, $50,000-74,999, $75,000-99,999, $100,000-149,999, $150,000-299,999, >$300,000. **B)** Distribution of total SD-WISE-7 scores (range 0 to 28). Mean (**SD**, standard deviation) was 18.5 (3.7), and median was 19 (interquartile range 16 to 21). **C)** Distributions of individual item scores (0 to 4), with means and standard error (**SE**). Self-Reflection, Decisiveness, Social Advising, and Pro-Social Behaviors were reverse-scored so that higher scores indicate greater wisdom. Percentages in panels B and C use the full analytic cohort (*N* = 161,775).

Total wisdom scores were calculated by summing responses to SD-WISE-7 items: 1) Emotional Regulation: *“I remain calm under pressure”*; 2) Self-Reflection: *“I avoid self-reflection”* (reverse coded, “**R**”); 3) Acceptance of Divergent Perspectives: *“I enjoy being exposed to diverse viewpoints”*; 4) Decisiveness: *“I tend to postpone making major decisions as long as I can”* (R); 5) Social Advising*: “I often don’t know what to tell people when they come to me for advice”* (R); 6) Spirituality: *“My spiritual belief gives me inner strength”*; 7) Pro-Social Behaviors: *“I avoid situations where I know my help will be needed”* (R). Likert scale options were coded numerically as follows: “Strongly disagree” = 0, “Disagree” = 1, “Neutral” = 2, “Agree” = 3, and “Strongly agree” = 4. See the **Methods** section for scoring and additional information. Wisdom items were mostly positively phenotypically inter-correlated after controlling for sex, age, education, and household income (*r_p-range_*= 0.01 ± 2.56E-03 to 0.32 ± 2.43E-03, false discovery rate [**FDR**] *p* < 0.001; **Supplementary Table 3**).

### SD-WISE-7 Items Phenotypically Correlate with 31 Demographics, Personality, Mental and Physical Health Measures

Total wisdom scores were FDR-significantly phenotypically correlated (***r_p_***) with all of the 31 demographic and SES variables, externalizing and impulsivity measures, and physical and psychiatric phenotypes tested (**Figure 2**; **Supplementary Table 4**).

**Figure 2.**
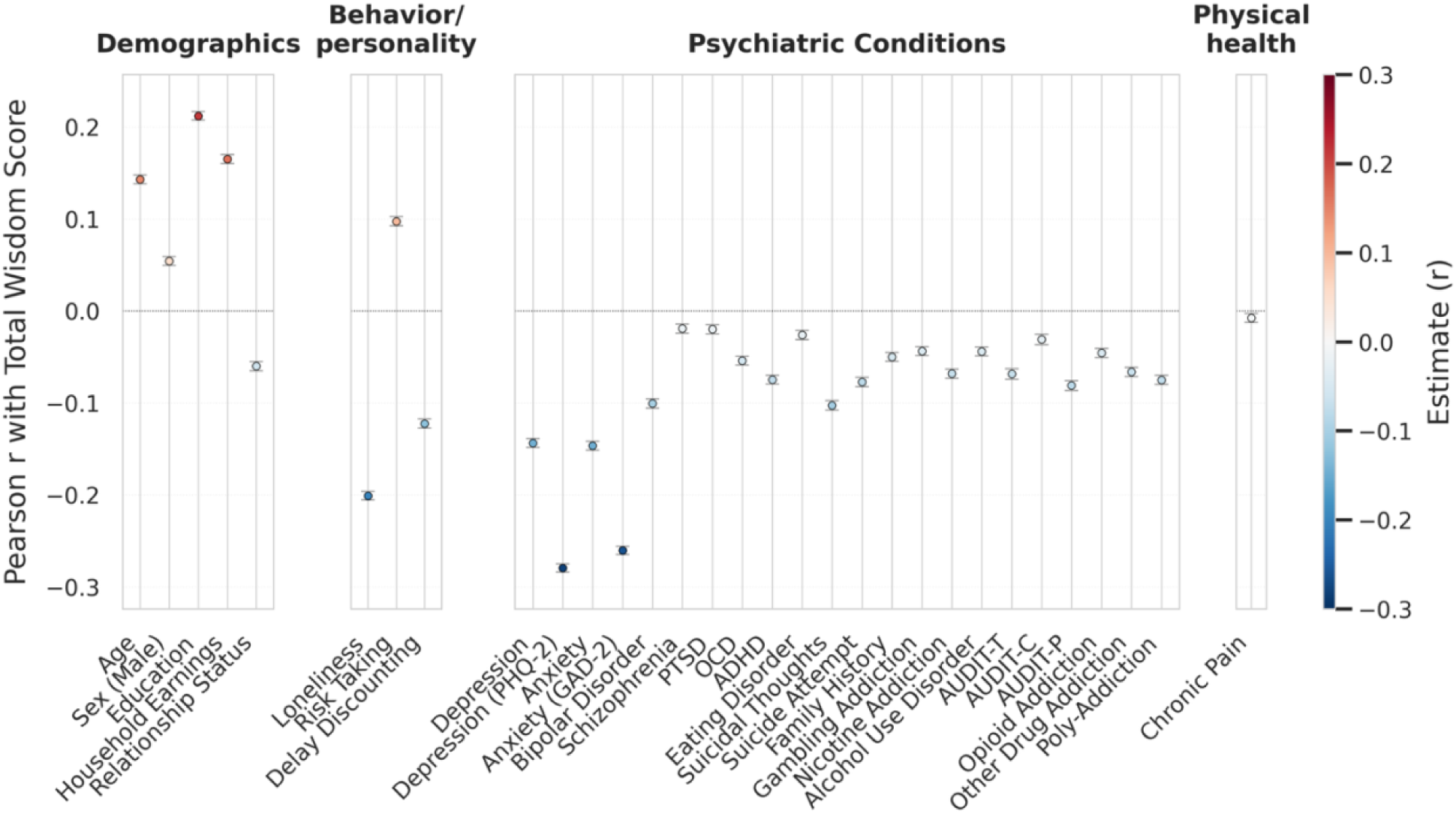
Phenotypic correlations. Unadjusted Pearson correlations (***r_p_***) between total wisdom scores and 31 traits, grouped by category. Points show the correlation estimate and bars the 95% confidence interval. Color denotes correlation direction and magnitude. All 31 phenotypes were significantly correlated with total wisdom scores after false discovery rate (**FDR**) correction across the 31 tests. See **Supplementary Table 4** for full statistics and **Supplementary Figure 1-2** for correlations among the SD-WISE-7 items. For sex correlations, male = 1 and female = 0.

For demographic and SES variables, total wisdom scores were positively correlated with older age (*r_p_* = 0.14 ± 2.46E-03, *p* < 5.00E-324), being male (*r_p_* = 0.05 ± 2.48E-03, *p* = 6.93E-106), greater levels of education (*r_p_* = 0.21 ± 2.43E-03, *p* < 5.00E-324), and higher household income (*r_p_* = 0.17 ± 2.52E-03, *p* < 5.00E-324). The correlations with older age and higher education were directionally consistent with prior findings, whereas the association with male sex was not consistent with previous findings reporting positive associations with female sex^6,7^.

Next, we examined correlations with externalizing-related traits. We observed a negative phenotypic correlation between total wisdom scores and alcohol consumption (**AUDIT-C**; *r_p_* = - 0.03 ± 2.93E-03, p = 4.90E-26), consistent with prior findings in adolescents^32^. We also identified a negative phenotypic correlation between total wisdom scores and delay discounting (i.e., the tendency to devalue rewards as their receipt is temporally delayed ^33^; *r_p_* = −0.12 ± 2.53E-03, *p* < 5.00E-324) and a positive phenotypic correlation with risk-taking (*r_p_* = 0.10 ± 2.48E-03, *p* < 5.00E-324).

For psychiatric traits and disorders, total wisdom scores showed negative phenotypic correlations directionally consistent with prior associations with wisdom measures, including loneliness (*r_p_* = −0.20 ± 2.44E-03, *p* < 5.00E-324)^7^, major depressive disorder (*r_p_* = −0.14 ± 2.46E-03, *p* < 5.00E-324) and depressive symptoms (i.e., PHQ-2 scores; *r_p_* = −0.28 ± 2.39E-03, *p* < 5.00E-324)^7^, anxiety disorder (*r_p_* = −0.15 ± 2.46E-03, *p* < 5.00E-324) and anxiety symptoms (i.e., GAD-2 scores; *r_p_* = −0.26 ± 2.40E-03, *p* < 5.00E-324)^7^, suicidality (i.e., suicidal thoughts, *r_p_* = −0.10 ± 2.66E-03, *p* = 2.96E-323; suicide attempts, *r_p_* = −0.08 ± 2.66E-03, *p* = 5.52E-185)^34^, and schizophrenia (*r_p_* = −0.02 ± 2.49E-03, *p* = 1.25E-14)^35^. We also identified negative phenotypic correlations with traits that had not been previously studied in relation to SD-WISE-7, including bipolar disorder (*r_p_* = −0.10 ± 2.48E-03, *p* < 5.00E-324), attention deficit hyperactivity disorder (**ADHD**; *r_p_* = −0.07 ± 2.48E-03, *p* = 9.88E-199), internalizing disorders (i.e., obsessive compulsive disorder *r_p_* = −0.05 ± 2.49E-03, *p* = 1.03E-104; eating disorders *r_p_* = −0.03 ± 2.49E-03, *p* = 5.31E-26; post-traumatic stress disorder [**PTSD**] *r_p_* = −0.02 ± 2.49E-03, *p* = 1.18E-15), and traits related to substance use disorders (e.g., multiple lifetime addictions, *r_p_* = −0.07 ± 2.48E-03, *p* = 6.34E-200; nicotine addiction, *r_p_* = −0.07 ± 2.48E-03, *p* = 1.72E-165; problematic consequences of alcohol use [i.e., **AUDIT-P** scores], *r_p_* = −0.08 ± 2.74E-03, *p* = 3.15E-192).

For physical health, we identified a negative phenotypic correlation between total wisdom scores and chronic pain (*r_p_* = −0.01 ± 2.49E-03, *p* = 2.16E-03).

After adjustment for sex, age, education, and household income, most phenotypic correlations remained significant after FDR correction, except for chronic pain, eating disorder, and schizophrenia (**Supplementary Table 4**).

### Genome-wide association study reveals an association between wisdom and a locus near *MANEA*

We performed GWAS on total wisdom scores in participants of European-like (*N_EUR_* = 131,870), Latin American-like (*N_LAT_* = 16,812), and African-like (*N_AFR_* = 4,702) genetic ancestries (**Supplementary Figures S3-S5**, **Supplementary Tables 5-7**). We identified a polygenic signal (χ^2^ = 1.07, 1.01, and 1.01; and λ_GC_ = 1.05, 1.00, and 1.00 for European-like, Latin American-like, and African-like groups, respectively) with minimal bias from population stratification (LDSC intercepts were 1.00 ± 0.01, 1.01 ± 0.01, and 1.02 ± 0.01, respectively). In the European-like cohort, total wisdom score SNP-based heritability (h^2^) was estimated to be 0.03 ± 0.005 (**Supplementary Table 8**). h^2^ estimates in the Latin American-like and African-like group were not significantly different from zero.

We then performed a multi-ancestry meta-analysis of 9,195,929 variants, combining ancestry-specific analyses totaling 153,384 participants (**Figure 3**). We identified a genome-wide significant locus on chromosome 6q16.1, led by rs2325631 (*p* = 1.41E-08; **Supplementary Table 9**; **Supplementary Figure 4-5**). This locus is near *MANEA*, which encodes α-endomannosidase, a glycosidase involved in N-linked oligosaccharide processing^36^. Prior studies have reported associations of *MANEA* variants with panic disorder and social anxiety disorder^37^, and with cocaine dependence and cocaine-induced paranoia^38^, though none of these variants are in linkage disequilibrium (**LD**) with rs2325631.

**Figure 3.**
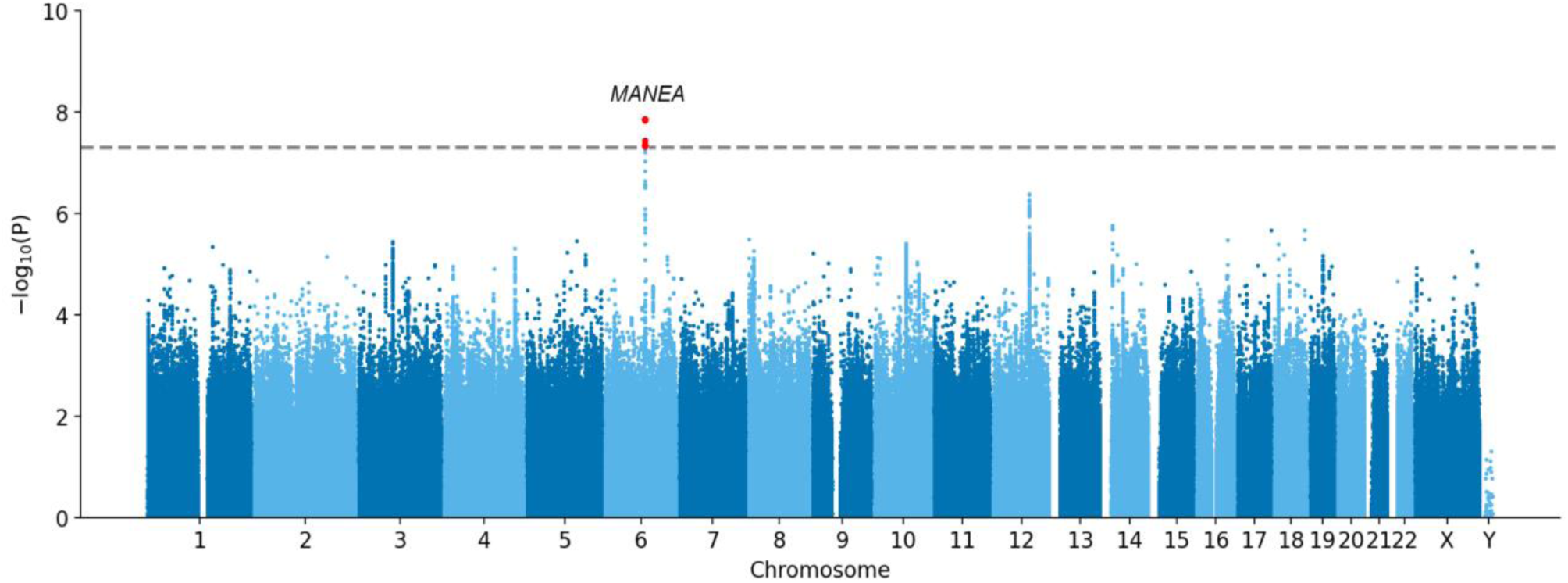
Multi-ancestry GWAS of wisdom. Manhattan plot of multi-ancestry GWAS meta-analysis results for total wisdom scores. Red dots indicate variants achieving genome-wide significance (*p* < 5.00E-08; dashed line) at one chromosome 6 locus near *MANEA*. See **Supplementary Table 5** for SNP-level quality control and filtering, and **Supplementary Figure 4** for the Q-Q plots.

To identify potential effector genes and pathways associated with total wisdom scores, we performed gene-based analyses (**MAGMA** and **H-MAGMA**), a transcriptome-wide association analysis (**S-PrediXcan**), and gene-set, tissue, and cell-type enrichment analyses (MAGMA; **Supplementary Tables 10-15**). MAGMA identified adenosine kinase (***ADK***) as the only gene significantly associated with total wisdom scores after Bonferroni correction across 19,576 genes (*p* = 2.43E-06). *ADK* encodes an enzyme that regulates adenosine concentrations^39^. This gene has been implicated in GWAS of body mass index^40–43^, dental development^44–46^, Alzheimer’s disease^47^, and drug experimentation^29^, among other traits. S-PrediXcan likewise identified *ADK* expression in the brain cerebellar hemisphere as the only significant gene-tissue association after correction within each tissue (*p* = 4.56E-07). H-MAGMA or MAGMA gene-set, tissue or cell-type enrichment analyses did not identify significant associations after their respective multiple-testing corrections.

### Wisdom is genetically correlated with 21 demographic, personality, externalizing, psychiatric, health, and cognitive traits

We examined genetic correlations (***r_g_***) between total wisdom scores and other demographic/SES, behavioral and personality, physical health, psychiatric, cognitive, and neuroimaging traits that have been measured in other cohorts (**Figure 4**; **Supplementary Figure 6**; **Supplementary Table 16**). These analyses were limited to the subset of European-like individuals because of the lack of well-powered GWAS available in the other ancestries. Traits were selected based on observed^6,7^ or theoretical phenotypic correlations with wisdom^13^. Of the 64 traits examined, we detail below the 21 traits that had significant genetic correlations with total wisdom scores.

**Figure 4.**
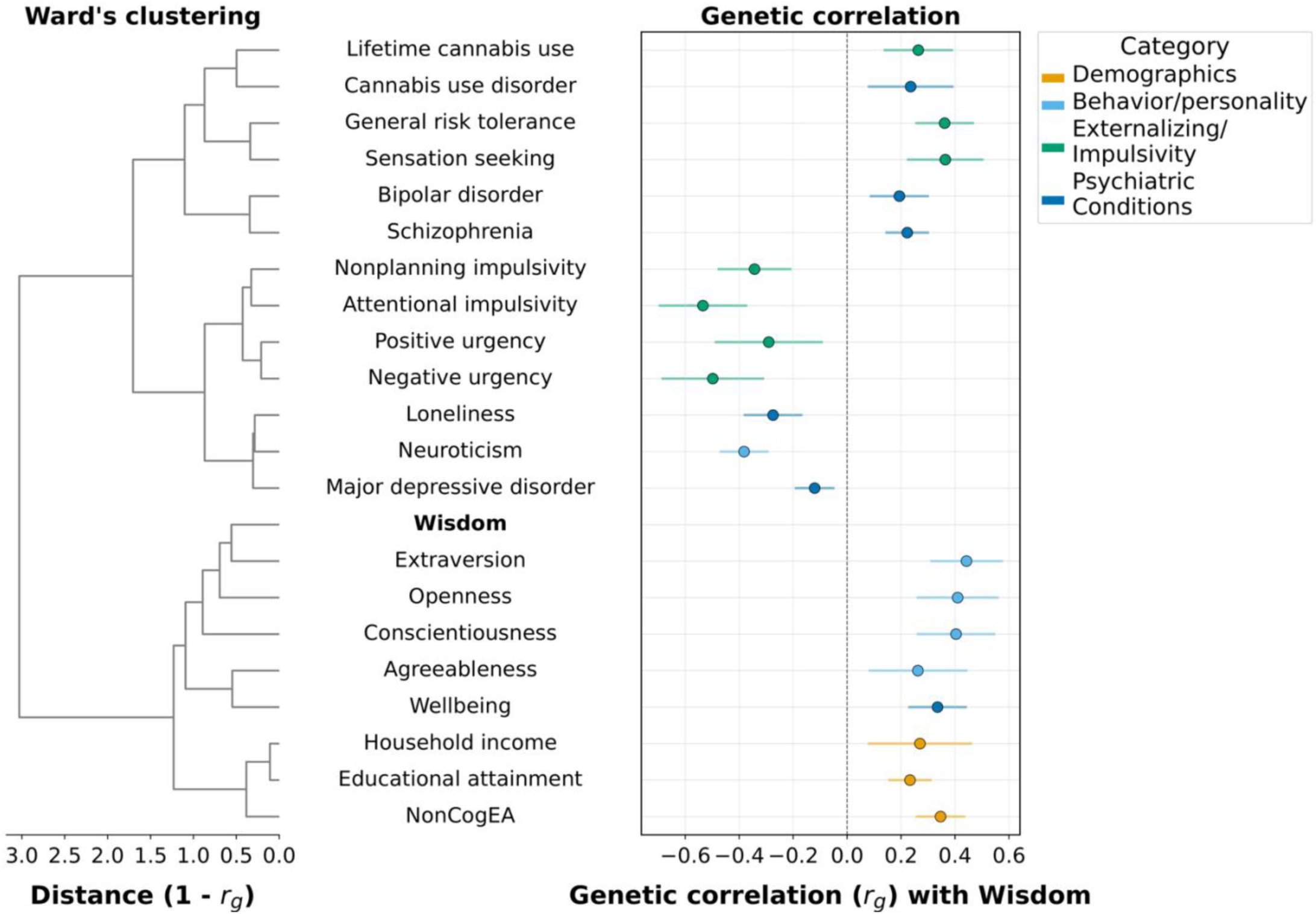
Genetic correlations between total wisdom scores and other selected traits. LDSC genetic correlations (***r_g_***) between total wisdom scores and traits spanning demographics, behaviour/personality, externalizing/impulsivity, psychiatric conditions, and physical health. Points show the *r_g_* estimate and horizontal bars the 95% confidence interval. Filled circles denote the 21 associations with Benjamini-Hochberg FDR *p* < 0.05 (computed across all 64 traits). Colours denote trait category. Rows are ordered by the dendrogram in the left panel, which shows Ward’s hierarchical clustering of the pairwise genetic correlations among these traits using 1 − *r_g_* as the distance metric; wisdom is included as a clustering leaf, and its row in the right panel is intentionally left blank. See **Supplementary Table 17** for statistics on all 64 traits.

#### Cluster analysis

We performed a cluster analysis restricted to traits significantly genetically correlated with wisdom. Wisdom was placed in a subcluster with several of the Big Five personality scales: extraversion, openness, conscientiousness, agreeableness, as well as wellbeing (**Supplementary Figure 7**). This personality cluster formed a supercluster with educational attainment, the non-cognitive component of educational attainment, and household income.

#### Demographics and Socioeconomic Status

Wisdom was genetically correlated with 3 out of the 6 demographic/SES indicator items tested. Consistent with SES phenotypic correlations, wisdom was positively genetically correlated with educational attainment (*r_g_* = 0.23 ± 0.04, *p* = 1.52E-08) and its non-cognitive component (**NonCog EA**; *r_g_* = 0.35 ± 0.05, *p* = 1.72E-13), and household income (*r_g_* = 0.27 ± 0.10, *p* = 6.20E-03), but not the Townsend Deprivation Index (*r_g_* = −0.03 ± 0.06, *p* = 0.69).

Wisdom was not genetically correlated with longevity (*r_g_* = −0.17 ± 0.12, *p* = 0.16) or parental lifespan (*r_g_ =* 0.02 ± 0.08, *p =* 0.77).

#### Personality

Wisdom was genetically correlated with 5 of the 6 tested personality traits. Among these were positive genetic correlations with the Big Five traits, namely extraversion (*r_g_* = 0.44 ± 0.07, *p* = 1.12E-10), openness (*r_g_* = 0.41 ± 0.08, *p* = 1.37E-07), conscientiousness (*r_g_* = 0.40 ± 0.07, *p* = 5.85E-08), and agreeableness (*r_g_* = 0.26 ± 0.09, *p* = 4.90E-03). Only one Big Five trait, neuroticism, was negatively genetically correlated with wisdom (*r_g_* = −0.38 ± 0.05, *p* = 1.87E-16).

#### Externalizing and Impulsivity

Wisdom was genetically correlated with 7 of the 15 externalizing and impulsivity traits tested. Total wisdom score was positively genetically correlated with general risk tolerance (*r_g_* = 0.36 ± 0.06, *p* = 6.94E-11), sensation seeking (*r_g_* = 0.36 ± 0.07, *p* = 5.22E-07), and cannabis lifetime use (*r_g_* = 0.26 ± 0.07, *p* = 5.90E-05), which was broadly consistent with phenotypic correlations. Negative genetic correlations included a subset of impulsivity dimensions: attentional impulsivity (*r_g_* = −0.53 ± 0.08, *p* = 1.89E-10), nonplanning impulsivity (*r_g_* = −0.34 ± 0.07, *p* = 9.02E-07), negative urgency (*r_g_* = −0.50 ± 0.10, *p* = 3.03E-07), and positive urgency (*r_g_* = −0.29 ± 0.10, *p* = 4.50E-03). Inconsistent with phenotypic correlations, wisdom was not genetically correlated with AUDIT-C (*r_g_* = 0.08 ± 0.06, *p* = 0.20), motor impulsivity (*r_g_* = −0.01 ± 0.08, *p* = 0.90), premeditation (*r_g_* = −0.12 ± 0.08, *p* = 0.16), perseverance (*r_g_* = −0.19 ± 0.10, *p* = 0.07), or with delay discounting after FDR correction (*r_g_* = −0.14 ± 0.07, *p* = 0.04).

#### Neurological Conditions

None of the 4 neurological conditions tested were genetically correlated with total wisdom score (*r_g-range_* = −0.06 ± 0.11 to 0.10 ± 0.05, *p_range_* = 0.06 to 0.79). (ADHD, *r_g_* = 0.10 ± 0.05, *p* = 0.06; autism spectrum disorder, *r_g_* = 0.08 ± 0.13, *p* = 0.53; Alzheimer’s, *r_g_* = −0.06 ± 0.11, *p* = 0.58;insomnia, *r_g_* = −0.02 ± 0.06, *p* = 0.79).

#### Psychiatric Conditions

Six out of 18 psychiatric disorders were genetically correlated with total wisdom score. Consistent with phenotypic correlations, wisdom was negatively genetically correlated with major depressive disorder (*r_g_* = −0.12 ± 0.04, *p* = 1.40E-03), and self-reported loneliness (*r_g_* = −0.27 ± 0.06, *p* = 8.45E-07). Wisdom scores were also positively genetically correlated with cannabis use disorder (*r_g_* = 0.24 ± 0.08, *p* = 3.80E-03), self-reported well-being (*r_g_* = 0.34 ± 0.06, *p* = 1.52E-09), bipolar disorder (*r_g_* = 0.19 ± 0.06, *p* = 5.00E-04), and schizophrenia (*r_g_* = 0.22 ± 0.04, *p* = 6.95E-08).

#### Physical Health

Despite previous reports relating wisdom to physical health^9,48^, only one of the 4 physical health traits we tested were genetically correlated with wisdom (type II diabetes, *r_g_*= 0.01 ± 0.05, *p* = 0.81; coronary artery disease, *r_g_* = 0.01 ± 0.04, *p* = 0.93; chronic pain, *r_g_ =* −0.12 ± 0.06, *p* = 0.03; physical illness factor, *r_g_ =* −0.04 ± 0.07, *p* = 0.55).

#### Cognition

Wisdom was not genetically correlated with any of the 3 cognitive traits tested: executive function (*r_g_* = −0.05 ± 0.05, *p* = 0.27), intelligence (*r_g_* = 0.04 ± 0.04, *p* = 0.39), or cognitive performance sourced from Demange et al. 2021 (*r_g_* = −0.01 ± 0.04, *p* = 0.83; **Supplementary Figure 7**).

#### Neuroimaging Measures

Wisdom was not genetically correlated with any of the 8 neuroimaging measures of brain volume examined (*r_g-range_* = −0.15 ± 0.12 to 0.22 ± 0.15, *p_range_*= 0.17 to 0.88).

### Wisdom polygenic scores were associated with health and well-being phenotypes but not cognitive performance in All of Us participants

To further explore the profile of wisdom’s genetic relationships with personality, health and well-being, and cognition, we examined the relationship between wisdom polygenic scores (**PGS**) and survey responses, electronic health records (**EHR**), and cognitive measures collected in AoU participants of European-like genetic ancestry (**Supplementary Table 17**).

#### Wisdom polygenic scores associate with self-reported wisdom proxies

We first examined PGS associations with 7 survey items that were exploratory proxies of the spirituality, pro-social behaviors, and emotional regulation items of SD-WISE-7 (*N_EUR_* ≤ 124,630; **Supplementary Table 18**). Wisdom PGS were associated with all 7 survey questions related to wisdom (e.g., *“How often do you find strength and comfort in your religion?“*, *“How often do you have someone to turn to for suggestions about how to deal with a personal problem?”, “In the last month, how often have you felt confident about your ability to handle your personal problems?”*; **Supplementary Table 18**) and explained up to 0.1% of their phenotypic variance (β*_range_* = 0.01 ± 2.93E-03 to 0.04 ± 3.84E-03, *p_range_* = 1.89E-30 to 1.99E-05, ΔR^2^*_range_* = 0.01 to 0.10%; **Supplementary Figure 8**).

#### Wisdom polygenic scores associate with self-reported measures of physical, mental, and social health, and well-being

We examined wisdom PGS associations with self-reported measures of health and well-being (*N* ≤ 211,029; **Figure 5**; **Supplementary Tables 19-21**). Broadly consistent with phenotypic and genetic correlations, wisdom PGS were associated with better self-reported ratings of social satisfaction (β = 0.03 ± 2.30E-03, *p* = 7.48E-32, PGS R^2^ = 0.06%), mental health (β = 0.02 ± 2.15E-03, *p* = 5.52E-27, PGS R^2^ = 0.05%), social functioning (β = 0.02 ± 2.17E-03, *p* = 1.14E-22, PGS R^2^ = 0.04%), quality of life (β = 0.02 ± 2.11E-03, *p* = 2.09E-19, PGS R^2^ = 0.04%), general health (β = 0.02 ± 2.15E-03, *p* = 7.32E-14, PGS R^2^ = 0.03%), physical health (β = 0.02 ± 2.18E-03, *p* = 1.08E-13, PGS R^2^ = 0.03%), and emotional health (β = 0.01 ± 2.18E-03, *p* = 3.24E-09, PGS R^2^ = 0.01%). In light of wisdom’s strong genetic correlations and genetic clustering with the Big Five personality traits and NonCogEA, we also examined if including these PGS (all five Big Five scores jointly) in the model impacted wisdom PGS associations (**Supplementary Tables 22-23**). Wisdom PGS associations were partially attenuated (β reductions of 17.70 to 29.47% for Big Five and 22.35 to 62.13% for NonCogEA) but still significantly associated with health and well-being measures when including PGS for the Big Five personality traits (β*_range_* = 0.01 ± 2.18E-03 to 0.02 ± 2.30E-03, *p_range_* = 4.26E-22 to 2.98E-05, PGS R^2^*_range_* = 0.01 to 0.04%) or NonCogEA in the model (β*_range_*= 0.01 ± 2.18E-03 to 0.02 ± 2.30E-03, *p_range_* = 8.86E-20 to 4.80E-03, PGS R^2^*_range_* = 3.21E-03 to 0.04%; **Supplementary Tables 22-23**). Consistent with the nonsignificant genetic correlation between total wisdom score and cognitive performance, we observed minimal change in wisdom PGS associations when cognitive performance PGS was included in the model (β*_range_* = 0.01-0.03 ± 2.18E-03 to 2.30E-03, *p_range_* = 8.76E-33 to 9.21E-10, PGS R^2^*_range_*= 0.02 to 0.07%; **Supplementary Table 24**).

**Figure 5.**
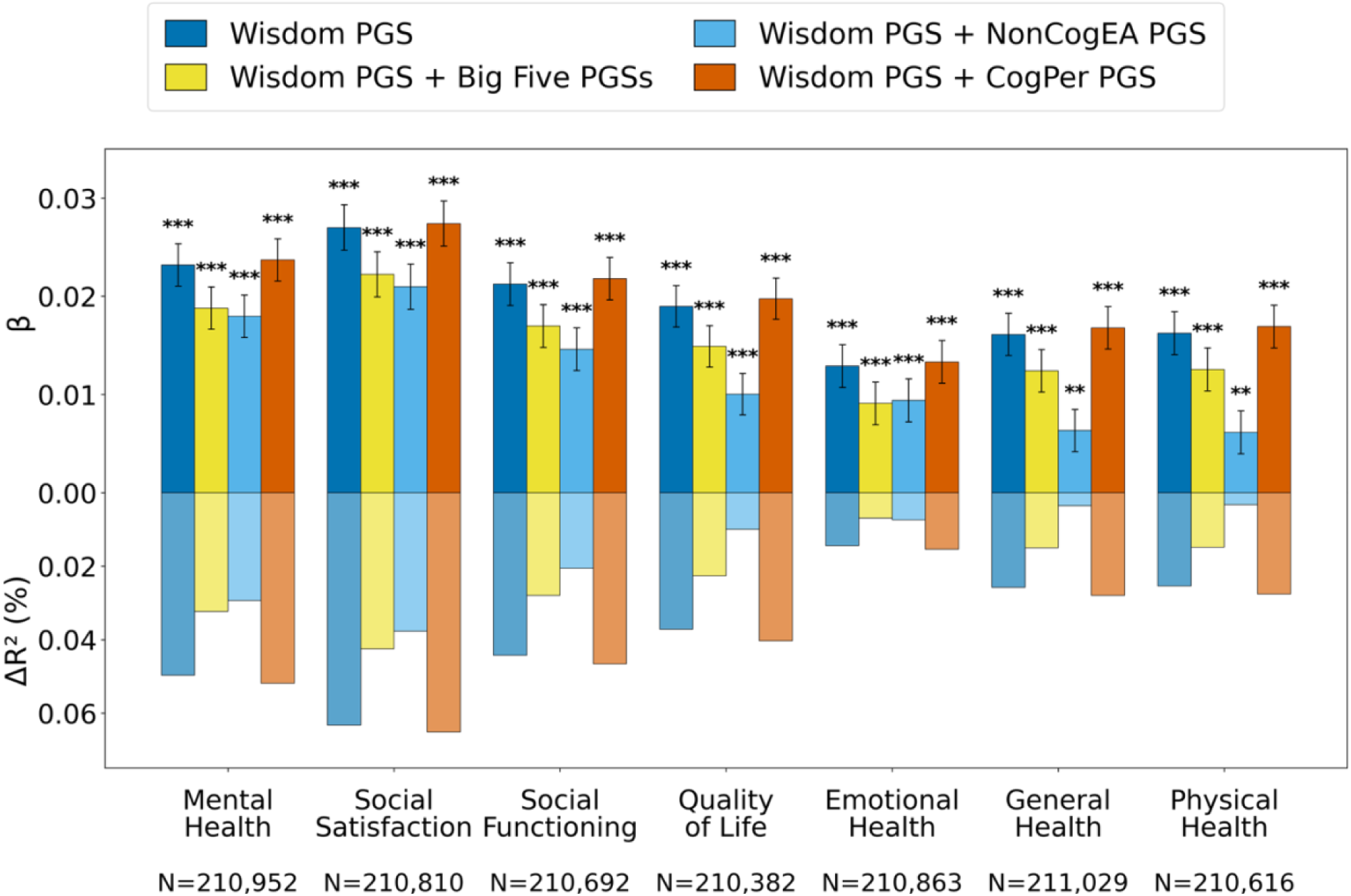
Wisdom PGS associations with measures of physical, mental, and social health, and well-being. Associations calculated in All of Us participants (*N_EUR_* < 211,030) across four models. Top bars show the effect (**β**), expressed in outcome units per standard deviation of wisdom PGS ± standard error. Bottom bars show variance explained (**ΔR²**) by wisdom PGS. Models include the main model (age, sex, PC1-10, SES, wisdom PGS); the main model plus all Big Five PGSs; the main model plus NonCogEA PGS; and the main model plus cognitive performance (**CogPer**) PGS. Asterisks denote FDR significance (\*\**q* < 0.01, \*\*\**q* < 0.001). See **Supplementary Tables 19-24** for model outputs.

Additionally, we examined wisdom PGS associations in a hypothesis-free manner with 1,600 medical outcomes through phenome-wide association study (**PheWAS**) of electronic health records (*N_EUR_* ≤ 155,512; **Supplementary Methods**; **Supplementary Figure 9**; **Supplementary Table 25**). We identified only one modest association surviving FDR correction, with ‘Chronic dermatitis due to solar radiation’ (OR = 1.06, 95% CI = 1.03 - 1.09, FDR *p* = 0.01).

#### Wisdom polygenic scores do not associate with cognitive performance measures

A subset of All of Us participants completed tasks that assess 1) impulsivity and decision making through delay discounting (*N_EUR_* = 14,733); 2) social cognition and emotional recognition through the Guess the Emotion task (*N_EUR_* = 17,960); 3) attention and inhibitory control through the Flanker task (*N_EUR_* = 14,185); and 4) sustained attention and cognitive control through the Gradual Onset Continuous Performance Task (*N_EUR_* = 14,734). We identified no associations between wisdom PGS and performance across any of the tasks (β*_range_* ± standard error = −5.07E-03 ± 7.68E-03 to 0.01 ± 8.37E-03, *p_range_*= 0.29 to 0.86; **Supplementary Table 26-28**). Associations remained insignificant when the Big Five, NonCogEA, or cognitive performance PGS were included in the model alongside the wisdom PGS (**Supplementary Figure 10**; **Supplementary Tables 29-31**).

## Discussion

This is the first study to explore the genetic basis of wisdom. We collected SD-WISE-7 responses at scale (N = 153,384) in 23andMe research participants for whom genome-wide genotype data were already available. We found that wisdom was heritable (h^2^SNP = 0.03), in alignment with other dispositional traits (e.g., ^24,27,49^). We identified one genome-wide significant locus, led by the intergenic variant rs2325631 near *MANEA*, and gene- and transcriptome-based analyses converged on *ADK*. Wisdom scores were associated with better health and well-being at phenotypic (within cohort) and genetic (with external cohorts) levels, supporting phenotypic findings by others^2,9^. Our genetic correlations were strongest with education and personality traits and not cognition. Wisdom PGS analysis similarly revealed positive associations with broad well-being outcomes that were partially attenuated by PGS of personality and non-cognitive educational attainment but not by cognitive performance. Together, our findings support a biological basis of wisdom and relationships with health and well-being that may be partially mediated by pleiotropy with personality traits that promote healthy lifestyles.

Wisdom is defined as a multifaceted personality trait with longstanding roots in spirituality that is composed of good decision-making abilities, flexible and reflective thinking, prosociality, emotional regulation, and the application of knowledge beyond cognitive ability^23^. SD-WISE-7 was developed to capture these characteristics^7^. Consistently, we found that wisdom was genetically correlated with other personality traits (e.g., the Big Five), but not at unity, emphasizing that wisdom is a distinct personality trait that is fundamental to human experiences.

We observed phenotypic and genetic associations between wisdom and numerous measures of physical and mental health, consistent with other personality traits (e.g., extraversion, openness, compassion, among others)^24,27,50–56^. Wisdom and its components (e.g., Pro-Social Behaviors, Emotional Regulation, and Social Advising) are malleable across the lifespan^18,19,57–60^, suggesting that wisdom may hold promise as a target for psychosocial and behavioral interventions to improve overall well-being^4^, as it has been explored for other personality traits^61^.

While we focused on total wisdom scores, divergences across the subcomponents indicate there is value to studying them individually. For example, although total wisdom scores were phenotypically correlated with affective measures, the items of Decisiveness (“*I tend to postpone making major decisions as long as I can*”) and Emotional Regulation (e.g., “*I remain calm under pressure*”) drove these associations. This is consistent with a prior SD-WISE study^9^, suggesting that Emotional Regulation may be a critical channel through which wisdom genetically relates to health. In addition, only examining total wisdom scores may occlude some associations. For example, older age showed strong but opposing item-level relationships (e.g., positive with Decisiveness, negative with Acceptance of Divergent Perspectives) that were masked by overall positive associations with total wisdom scores. Whether these item-specific patterns extend to genetic correlations remains unknown, as item-level GWAS data were unavailable.

Some conceptions of wisdom include intelligence, reasoning, and knowledge among its components^62^. Modern psychosocial frameworks and study measures, however, conceptualize wisdom as distinct from intelligence^3,63^. In particular for SD-WISE, wisdom scores are dissociable from fluid intelligence, executive function, and processing speed^4^. This is consistent with our findings that wisdom scores are genetically uncorrelated with intelligence/cognitive function, executive function, and cognitive performance, and wisdom PGS associations were unaltered when modelled alongside cognitive performance PGS. Wisdom PGS also did not associate with the four cognitive tasks collected in All of Us participants, despite expectations that some tasks (e.g., emotional recognition) would reflect SD-WISE-7 item responses. While this may suggest no associations between wisdom score and broad cognitive function, another study has identified modest positive associations between performance-based wisdom measures and crystallized intelligence^64^. We cannot rule out wisdom associations with cognition could be overlooked by our use of the SD-WISE-7 self-report and/or relatively small target samples.

Despite the lack of associations with cognition, wisdom was phenotypically and genetically correlated with educational attainment. Many factors outside of cognitive ability contribute to educational attainment, including socioeconomic circumstance^65^ and personality^66,67^. We found that the non-cognitive genetic component of educational attainment (**NonCogEA**)^68^ was genetically correlated with total wisdom scores, and wisdom PGS associations were attenuated when modeled jointly with NonCogEA PGS, suggesting substantial shared genetic influences between the two traits. We suggest that wisdom’s genetic relationship with educational attainment is sustained by non-cognitive factors (e.g., personality, SES) that support higher achievement, rather than with cognitive ability.

Overall, we find that wisdom phenotypically and genetically associates with better physical and mental health, consistent with prior phenotypic observations^1,7,9^. This pattern was concentrated in psychiatric rather than physical health, though we tested far fewer physical health traits. Perhaps surprisingly, wisdom was not genetically correlated with parental lifespan or longevity, despite the theory that wisdom accrued over life promotes post-reproductive inclusive fitness^22,23^. Instead, we found that wisdom PGS were most strongly associated with self-reported measures of social satisfaction and mental health, compared to psychiatric diagnosis from electronic health records, suggesting that wisdom’s genetic signal tracks broader psychosocial outcomes.

While we generally found favorable wisdom associations with health, we report some unexpected genetic relationships with schizophrenia, bipolar disorder, and cannabis use traits. Total wisdom scores were negatively phenotypically correlated with schizophrenia and bipolar disorder, whereas genetic correlations were positive. Furthermore, although we had no phenotypic measures for cannabis use, genetic correlations with lifetime cannabis use and cannabis use disorder were also positive despite finding no other genetic correlations with substance use. This is not unprecedented: favourable genetic relationships between schizophrenia and educational attainment^69^, or alcohol use and psychiatric conditions^70^, were explained by their shared genetic liability with other unaccounted for traits (e.g., SES). Likewise, other personality traits (e.g., openness, extraversion), which also show similar paradoxical positive genetic correlations with schizophrenia and cannabis use, may mediate the inconsistent observations in our study (**Supplementary Figure 6**).

This study has several limitations. First, total wisdom scores were quantified through subjective self-reported items. SD-WISE-7 is the most feasible wisdom measure to use at the scale required for GWAS^7^, using structural equation modeling to weight each of the SD-WISE-7 items (rather than using a sum of the scores) may better fit our data (**Supplementary Figure 11**). Furthermore, wisdom could be defined in other ways, including with other questionnaires, which might yield different results than those presented here. Second, while social desirability may influence participant self-reports, one previous study found a low correlation between SD-WISE responding and social desirability^6^. Third, wisdom measures were collected as part of a larger study including individuals who all reported using opioids in their lifetime, which could have biased our observations. However, we did not find any evidence that opioid use was genetically correlated with total wisdom scores. Fourth, given the absence of clear functional evidence linking the *MANEA* locus to wisdom and the pleiotropic associations of *ADK* (e.g., ^41,45^), both findings require replication. Fifth, our analyses were all cross-sectional and underpowered to study the direction or causality of wisdom relationships. Finally, only the European-like ancestry GWAS was well-powered to detect specific associations, which may be particularly relevant to wisdom given there are known cross-cultural differences in wisdom perception^71^.

Overall, we present the first evidence that wisdom is a heritable trait with genetic links to personality, behavior, and health, but not cognition. Our results show that greater genetic liability to wisdom is associated with a constellation of health-related outcomes that are only partially attributable to the genetics of other personality dimensions. As a biologically-grounded yet environmentally malleable trait, future investigations with better powered genetic data may provide additional support for wisdom as a valuable target to broadly improve well-being.

## Methods

### Participants and SD-WISE-7 Assessment

Participants included research-consented individuals from 23andMe Research Institute, a nonprofit medical research organization. As part of a larger survey including 198,906 genotyped research-consented participants who endorsed using opioids in their lifetime^30^, participants completed the SD-WISE-7 wisdom questionnaire^7,31^. The data included in this study was collected from May 23, 2023 to May 15, 2025 (recruitment is still ongoing). Recruitment targeted US-based 23andMe participants who had previously reported taking a prescription pain medication. Participants were invited via email and could access the survey through the 23andMe website and app. No compensation was provided. Participants provided informed consent and volunteered to participate in the research online under a protocol approved by the external AAHRPP-accredited Salus IRB (https://www.versiticlinicaltrials.org/salusirb).

We evaluated wisdom with SD-WISE-7^7^. Participants rated items on a five-point Likert scale with responses including “Strongly disagree”, “Disagree”, “Neutral”, “Agree”, and “Strongly agree”. Responses were scored 0-4; the four negatively worded items were reverse coded (denoted below as “R”). Total wisdom scores were the sum of all seven items and ranged 0 −28. Complete responses to all seven items were required. The 7 survey items were:

1) Emotional Regulation: I remain calm under pressure.
2) Self-Reflection: I avoid self-reflection (R).
3) Acceptance of Divergent Perspectives: I enjoy being exposed to diverse viewpoints.
4) Decisiveness: I tend to postpone making major decisions as long as I can (R).
5) Social Advising: I often don’t know what to tell people when they come to me for advice (R).
6) Spirituality: My spiritual belief gives me inner strength.
7) Pro-Social Behaviors: I avoid situations where I know my help will be needed (R).

### Phenotypic Correlations

Phenotypic correlations (*r_p_*) of total wisdom scores and individual SD-WISE-7 items with 31 demographic, health-related behavioral measures, and psychiatric phenotypes were calculated in up to 161,775 research participants with complete data available. We calculated unadjusted Pearson correlations and partial correlations that were adjusted for age, sex, education, and household income. Multiple testing was accounted for by applying a FDR correction across the 31 tests in each model. See **Supplementary Materials** for additional details.

### Genotyping and Genome-wide Association Study

Ancestry-specific GWAS included 131,870 European-like, 16,812 Latin American-like, and 4,702 African-like unrelated participants. Ancestry was determined through an analysis of local ancestry (**Supplementary Materials**). DNA extraction and genotyping were performed on saliva samples at clinical laboratories of Laboratory Corporation of America, certified by the Clinical Laboratory Improvement Amendments (**CLIA**) and accredited by the College of American Pathologists (**CAP**). Quality control, imputation, and genome-wide analysis were conducted by 23andMe (**Supplementary Materials**), as previously described^72–76^. Age, sex, ancestral-specific genotype principal components (five for European-like, nine for Latin American-like, six for African-like groups), and genotype-platform indicators were included as covariates. Association tests were conducted via linear regression under an additive model using a proprietary pipeline developed internally by 23andMe (*p* < 5.00E-08). Between 15,700,769 and 32,738,086 variants passed the original 23andMe quality control (**Supplementary Table 5**). Previous associations with the SNPs implicated in our GWAS were examined with GWAS Catalog^77^. Credible sets were calculated by first calculating Wakefield’s Approximate Bayes Factor for each SNP and assuming a prior variance (**W**) of 0.1. The credible set was then estimated from the Approximate Bayes Factor as previously described^78^. Additional QC was applied with the EasyQC pipeline^79^.

### GWAS Meta-Analysis

METAL^80^ was used to meta-analyze ancestry-specific GWAS. A total of 9,195,929 SNPs were included in the meta-analysis. We used the fixed-effect inverse-variance-weighted method assuming shared risk effects across ancestries. Inflation of test statistics due to polygenicity or cryptic relatedness was assessed using the LDSC attenuation ratio ((LDSC intercept − 1)/(mean of association chi-square statistics − 1)). Independent loci were defined across SNPs with *p* < 5.00 E-08 and Linkage Disequilibrium (**LD**) *r*^2^ < 0.1 within a 1 Mb window, based on the structure of the Haplotype Reference Consortium (**HRC**) multi-ancestry reference panel for the multi-ancestry meta-analysis. Cohort heterogeneity was examined with the *I*^2^ inconsistency metric. Risk loci and lead SNPs were identified with PLINK v2.0^81^ using a range of 3 Mb, *r*^2^ > 0.1 and the 1000 Genomes Phase 3 European reference panel for both EUR and meta-analysis^82^. Genomic risk loci that were located < 1 Mb apart were incorporated into a single locus. Several variants were contained in the locus identified; we used COJO in GCTA^83^ to define independent variants by conditioning them on the most significant variant within the locus and identify independent associations (*p* < 5.00E-08).

### Functional Annotation and Gene-Based Analyses

#### MAGMA gene-based, pathway, and tissue analyses

We used Multi-marker Analysis of GenoMic Annotation (**MAGMA**, v1.10, Ensembl v110; ^84^) to annotate total wisdom score-associated SNPs to protein-coding genes and perform SNPwise-mean gene-based tests. LD was estimated using the 1000 Genomes European reference sample, and significance determined following correction for the number of gene associations tested (*N_genes_* = 19,576; Bonferroni *p <* 2.55E-06). Using the MAGMA option in the web-based platform Functional Mapping and Annotation of Genome-Wide Association Studies (**FUMA** v1.8.3; ^85^), we explored total wisdom score genetic associations with gene sets and gene expression profiles across tissue types. Gene set analysis was conducted on 17,011 gene sets (Bonferroni *p* < 2.94E-06) and gene ontology (**GO**) terms curated from the Molecular Signatures Database (MsigDB v7.0). Tissue-specific gene expression profiles were assessed in 54 tissue types (Bonferroni *p <* 9.26E-04) and 30 general tissue types (Bonferroni *p* < 1.67E-03) with expression in each tissue tested while conditioning on average expression across tissues. Using Genome-Tissue Expression (**GTEx**, v8) RNA-seq data, gene expression values were log_2_ transformed from the average Reads Per Kilobase Million (max value = 50) per tissue.

#### S-PrediXcan

We performed a transcriptome-wide association study using S-PrediXcan (v0.7.5; ^86^) to identify eQTL-linked genes associated with total wisdom scores. S-PrediXcan uses genetic information to predict gene expression levels and test eQTLs correlations with total wisdom scores across 13 tissues (*N_genes_* per tissue = 2,549 to 6,775; Bonferroni *p* < 7.38E-06 to 1.96E-05). S-PrediXcan uses precomputed tissue weights from the GTEx project database (https://www.gtexportal.org/) as the reference transcriptome dataset via Elastic net models. Input data included summary statistics, transcriptome tissue data, and covariance matrices of the SNPs within each gene model (HapMap SNP set available at the PredictDB Data Repository; https://predictdb.org/categories/downloads/) from all available tissues.

### LDSC heritability and genetic correlations

Linkage Disequilibrium Score regression (**LDSC**; https://github.com/bulik/ldsc) was used to calculate SNP-based heritability (***h2_SNP_***) and genetic correlations (***r_g_***; ^87,88^). *h2_SNP_* was calculated using pre-computed LD scores suitable for each ancestry (EUR: “eur_w_ld_chr/”; LAT and AFR were calculated locally using PLINK v2.0). Using EUR data, cross-trait *r_g_* were calculated between total wisdom scores and publicly available GWAS summary statistics for 64 demographic, health, psychiatric, cognitive, neuroimaging, and personality traits (**Supplementary Methods**). Benjamini-Hochberg FDR *correction* was applied across the 64 wisdom-trait tests (*q* < 0.05).

### Polygenic score analyses

#### All of Us participants

We tested the associations between wisdom PGS with select survey, EHR, or cognitive test data available in the US-based All of Us Research Program (**AoU**; v8 release; *N* < 230,057). See publications from the AoU Research Program for details on recruitment, study measures, and genomic data^89–91^. Unrelated participants (kinship score estimation ≤ 0.1, provided by AoU) who were assigned male or female at birth, had applicable phenotypic data, had whole genome sequencing (**WGS**) data, and whose genomes were statistically correlated with the genomes of European-like reference populations based on PCs provided by AoU^90^ were included in the study.

#### Wisdom Proxies, Health, Well-Being, and Cognitive Phenotypes

We could not directly evaluate the associative validity of wisdom PGS on a wisdom phenotype because no large-scale cohorts with genetic data that evaluate wisdom measures. Instead, seven survey questions were selected from the AoU Social Factors of Health survey as self-reported proxies to SD-WISE-7 items 1, 5, and 6 (*N* < 134,464). There were no sufficient proxies for the remaining SD-WISE-7 items. Seven items from the Overall Health survey were selected as measures of self-reported physical, mental, emotional, and social health, as well as overall health and quality of life (*N* < 230,057). See **Supplementary Table 17** for concept IDs, full questions, and additional information. We also tested total wisdom score PGS associations with task-based cognitive performance. This included the gradual continuous performance task (i.e., City or Mountain) with discrimination ability (*dprime*) being the primary response measure; the delay discounting task (i.e., Now or Later) with the natural log of the average final discounting factors (*lnk*) being the primary response measure; the flanker task (i.e., Left or Right) with rate-correct score (*rcs_interference*) being the primary response measure; and a variant of the facial recognition task (i.e., Guess the Emotion) with the proportion of correct responses (*accuracy*) being the primary response measure. For each cognitive task, one record with a nonmissing outcome per participant was retained, selecting the first such record in input-file order. PGS associations were determined through linear regression and adjusted for participant age, sex, the first 10 PCs, and zipcode-based socioeconomic deprivation index (**ZSDI**).

#### Polygenic score calculation and linear regression analysis

Associations between wisdom PGS and AoU phenotypes were conducted in those of European-like genetic ancestry due to limitations in power and that PGS are poorly portable across ancestries^92^. We used the Allele Count/Allele Frequency (**ACAF**) threshold SNP callset curated by AoU (minor allele frequency [**MAF**] > 1% or allele counts over 100). Using PRS-CS “auto” v1.1.0^93^, the SNP callset was filtered to biallelic SNPs present in the HapMap3 European ancestry set. PGS were created from 989,284 weighted SNPs using the score function in PLINK (v1.9)^81^. The OLS function in the Python package *statsmodels* was used to fit linear regression models for each continuous trait modeled with PGS(s) and covariates. Covariates included age, sex, the first 10 global PCs provided by AoU, and ZSDI. For the continuous-trait analyses, PGSs were standardized within the QC-filtered European-like ancestry cohort; cognitive outcomes were standardized within each task-specific analysis sample, whereas survey outcomes retained their encoded scales. Adjusted R^2^ values were obtained from the fitted linear regression models. Wisdom PGS ΔR^2^ was calculated by subtracting the adjusted R² of the null model excluding wisdom PGS from the adjusted R² of the model including wisdom PGS, using the same participants and retaining the same covariates and other PGSs, where applicable. Benjamini-Hochberg FDR correction was applied across the seven wisdom-proxy outcomes, across the seven health and well-being outcomes separately within each model, and across the four cognitive tasks separately within each model.

## Data Availability

We will provide 23andMe summary statistics for the top clumped 10,000 SNPs upon publication. Full GWAS summary statistics will be made available through 23andMe to qualified researchers under an agreement with 23andMe that protects the privacy of the 23andMe participants. Please visit https://research.23andme.com/collaborate/#dataset-access/ for more information and to apply to access the data.

We will share the Jupyter notebooks used for PGS analysis in AoU with registered AoU researchers upon request.

## Acknowledgements

HHAT is funded by a Canadian Institute of Health Research (CIHR) Postdoctoral Fellowship (#187919). VT, SS, ER, and the 23andMe Research Team are employed by the 23andM Research Institute. AAP is supported by NIDA (P50DA037844). SSR is funded through the National Institute on Drug Abuse (NIDA DP1DA054394). JG, JJM, AAP and SSR are funded by R01DA061977. LVR is funded by the Tobacco-Related Disease Research Program (TRDRP T35FT9589). NCK is funded by the TRDRP (T33KT6694) and the National Institute on Alcohol Abuse and Alcoholism grant number (L40AA031140). This work was partially funded by a Wisdom Pilot Grant Award from the Stein Institute and Center for Healthy Aging at UC San Diego.

We would like to thank Drs. Ellen Lee and Dilip Jeste for their guidance in selecting the wisdom questionnaire and for their contributions to the initial conceptualization of the research.

We would like to thank the research participants and employees of 23andMe for making this work possible. Participants provided informed consent and volunteered to participate in the research online, under a protocol approved by the external AAHRPP-accredited Institutional Review Board (**IRB**), Ethical & Independent (**E&I**) Review Services. As of 2022, E&I Review Services is part of Salus IRB (https://www.versiticlinicaltrials.org/salusirb). The following members of the 23andMe Research Team contributed to this study: Noura S. Abul-Husn, Aditya Ambati, Stella Aslibekyan, James R. Ashenhurst, Adam Auton, Robert K. Bell, Rebecca M. K. Berns, Kahsaia de Brito, Stacey B. Detweiler, Emily DelloRusso, Sayantan Das, William A. Freyman, Chris German, Anna Guan, Éadaoin Harney, Alejandro Hernandez-Chavez, Barry Hicks, David A. Hinds, Ethan M. Jewett, Bertram L. Koelsch, Matthew J. Kmiecik, Alan Kwong, Katelyn Kukar Bond, Alisa P. Lehman, Keng-Han Lin, Matthew H. McIntyre, Steven J. Micheletti, Alexander Moran, G. David Poznik, Emily M. Rios, Shubham Saini, Anjali J. Shastri, Qiaojuan Jane Su, Ruth I. Tennen, Vinh Tran, Joyce Y. Tung, Catherine H. Weldon, Peter R. Wilton, R. Ryanne Wu, Jingran Wen, Wanwan Xu.

We would also like to thank The Externalizing Consortium for sharing the GWAS summary statistics of externalizing. The Externalizing Consortium: Principal Investigators: Danielle M. Dick, Philipp Koellinger, K. Paige Harden, Abraham A. Palmer. Lead Analysts: Richard Karlsson Linnér, Travis T. Mallard, Peter B. Barr, Sandra Sanchez-Roige. Significant Contributors: Irwin D. Waldman. The Externalizing Consortium has been supported by the National Institute on Alcohol Abuse and Alcoholism (R01AA015416 -administrative supplement), and the National Institute on Drug Abuse (R01DA050721). Additional funding for investigator effort has been provided by K02AA018755, U10AA008401, P50AA022537, as well as a European Research Council Consolidator Grant (647648 EdGe to Koellinger). The content is solely the responsibility of the authors and does not necessarily represent the official views of the above funding bodies. The Externalizing Consortium would like to thank the following groups for making the research possible: 23andMe, Add Health, Vanderbilt University Medical Center’s BioVU, Collaborative Study on the Genetics of Alcoholism (COGA), the Psychiatric Genomics Consortium’s Substance Use Disorders working group, UK10K Consortium, UK Biobank, and Philadelphia Neurodevelopmental Cohort.

We gratefully acknowledge *All of Us* participants for their contributions, without whom this research would not have been possible. We also thank the National Institutes of Health’s *All of Us* Research Program for making available the participant data examined in this study. The *All of Us* Research Program is supported by the National Institutes of Health, Office of the Director: Regional Medical Centers: 1 OT2 OD026549; 1 OT2 OD026554; 1 OT2 OD026557; 1 OT2 OD026556; 1 OT2 OD026550; 1 OT2 OD 026552; 1 OT2 OD026553; 1 OT2 OD026548; 1 OT2 OD026551; 1 OT2 OD026555; IAA #: AOD 16037; Federally Qualified Health Centers: HHSN 263201600085U; Data and Research Center: 5 U2C OD023196; Biobank: 1 U24 OD023121; The Participant Center: U24 OD023176; Participant Technology Systems Center: 1 U24 OD023163; Communications and Engagement: 3 OT2 OD023205; 3 OT2 OD023206; and Community Partners: 1 OT2 OD025277; 3 OT2 OD025315; 1 OT2 OD025337; 1 OT2 OD025276.

## Author Contributions

SSR and AAP conceived the idea. FM, HHAT, JG, JJM, and LVR conducted the analyses. FM, HHAT and SSR wrote the manuscript. FM created the visualizations. NS and DG provided consultation services. VT, SS, and ER contributed formal analyses and curation of 23andMe data. All authors reviewed and approved the manuscript.

## References

1. Ardelt, M. Empirical Assessment of a Three-Dimensional Wisdom Scale. Res Aging 25, 275–324 (2003).

2. Jeste, D. V. & Lee, E. E. The Emerging Empirical Science of Wisdom: Definition, Measurement, Neurobiology, Longevity, and Interventions. Harv Rev Psychiatry 27, 127–140 (2019).

3. Jeste, D. V. et al. Expert consensus on characteristics of wisdom: a Delphi method study. Gerontologist 50, 668–680 (2010).

4. Lindbergh, C. A. et al. Wisdom and fluid intelligence are dissociable in healthy older adults. Int Psychogeriatr 34, 229–239 (2022).

5. Nisbett, R. E. et al. Intelligence: new findings and theoretical developments. Am Psychol 67, 130–159 (2012).

6. Thomas, M. L. et al. A new scale for assessing wisdom based on common domains and a neurobiological model: The San Diego Wisdom Scale (SD-WISE). J Psychiatr Res 108, 40–47 (2019).

7. Thomas, M. L. et al. Abbreviated San Diego Wisdom Scale (SD-WISE-7) and Jeste-Thomas Wisdom Index (JTWI). Int Psychogeriatr 34, 617–626 (2022).

8. Webster, J. D., Westerhof, G. J. & Bohlmeijer, E. T. Wisdom and mental health across the lifespan. J Gerontol B Psychol Sci Soc Sci 69, 209–218 (2014).

9. Jeste, D. V. et al. Is spirituality a component of wisdom? Study of 1,786 adults using expanded San Diego Wisdom Scale (Jeste-Thomas Wisdom Index). J Psychiatr Res 132, 174–181 (2021).

10. Bergsma, A. & Ardelt, M. Self-Reported Wisdom and Happiness: An Empirical Investigation. J Happiness Stud 13, 481–499 (2012).

11. Ardelt, M. Wisdom and life satisfaction in old age. J Gerontol B Psychol Sci Soc Sci 52B, P15–P27 (1997).

12. Lee, E. E. et al. High prevalence and adverse health effects of loneliness in community-dwelling adults across the lifespan: role of wisdom as a protective factor. Int Psychogeriatr 31, 1447–1462 (2019).

13. Meeks, T. W. & Jeste, D. V. Neurobiology of wisdom: a literature overview. Arch Gen Psychiatry 66, 355–365 (2009).

14. Bang, J., Spina, S. & Miller, B. L. Frontotemporal dementia. Lancet 386, 1672–1682 (2015).

15. Rogers, K., Dziobek, I., Hassenstab, J., Wolf, O. T. & Convit, A. Who cares? Revisiting empathy in Asperger syndrome. J Autism Dev Disord 37, 709–715 (2007).

16. Wink, P. & Helson, R. Practical and transcendent wisdom: Their nature and some longitudinal findings. J Adult Dev 4, 1–15 (1997).

17. Wang, Y. & Cheng, H. Longitudinal Association Between Mindfulness and Wisdom: A Follow-Up Study in Emerging Adulthood. J Intell 13, 122 (2025).

18. Lee, E. E. et al. Outcomes of Randomized Clinical Trials of Interventions to Enhance Social, Emotional, and Spiritual Components of Wisdom: A Systematic Review and Meta-analysis. JAMA Psychiatry 77, 925–935 (2020).

19. Treichler, E. B. H. et al. A pragmatic trial of a group intervention in senior housing communities to increase resilience. Int Psychogeriatr 32, 173–182 (2020).

20. Al-Refae, M., Al-Refae, A., Munroe, M., Sardella, N. A. & Ferrari, M. A Self-Compassion and Mindfulness-Based Cognitive Mobile Intervention (Serene) for Depression, Anxiety, and Stress: Promoting Adaptive Emotional Regulation and Wisdom. Front Psychol 12, 648087 (2021).

21. Hawkes, K. Grandmothers and the evolution of human longevity. Am J Hum Biol 15, 380–400 (2003).

22. Greve, W. & Bjorklund, D. F. The Nestor effect: Extending evolutionary developmental psychology to a lifespan perspective. Developmental Review 29, 163–179 (2009).

23. Jeste, D. V. et al. The New Science of Practical Wisdom. Perspect Biol Med 62, 216–236 (2019).

24. Okbay, A. et al. Genetic variants associated with subjective well-being, depressive symptoms, and neuroticism identified through genome-wide analyses. Nat Genet 48, 624–633 (2016).

25. Boardman, J. D., Blalock, C. L. & Button, T. M. M. Sex differences in the heritability of resilience. Twin Res Hum Genet 11, 12–27 (2008).

26. Abdellaoui, A. et al. Phenome-wide investigation of health outcomes associated with genetic predisposition to loneliness. Hum Mol Genet 28, 3853–3865 (2019).

27. Schwaba, T. et al. Robust inference and correlates from genetic associations with personality. Nature 10.1038/s41586-026-10992-9 (2026).

28. Day, F. R., Ong, K. K. & Perry, J. R. B. Elucidating the genetic basis of social interaction and isolation. Nat Commun 9, 2457 (2018).

29. Sanchez-Roige, S. et al. Genome-Wide Association Studies of Impulsive Personality Traits (BIS-11 and UPPS-P) and Drug Experimentation in up to 22,861 Adult Research Participants Identify Loci in the CACNA1I and CADM2 genes. J Neurosci 39, 2562–2572 (2019).

30. Gonzalez, J. et al. How it begins: Initial response to opioids strongly predicts self-reported opioid use disorder. Preprint at 10.1101/2025.03.21.25324409 (2025).

31. Courchesne-Krak, N. S. et al. Prescription Opioid Medication Survey: A Tool to Collect Deep Phenotypic Data on the Multifactorial Pathways to Opioid Use Disorder in Clinical and Population-Based Cohorts. Complex Psychiatry 11, 72–93 (2025).

32. Perry, C. L. et al. The Measurement of Wisdom and Its Relationship to Adolescent Substance Use and Problem Behaviors. Journal of Child & Adolescent Substance Abuse 12, 45–63 (2002).

33. Bickel, W. K. et al. Excessive discounting of delayed reinforcers as a trans-disease process: Update on the state of the science. Curr Opin Psychol 30, 59–64 (2019).

34. Dewangan, R. L., Gupta, S., Jeste, D. V., Longkumer, I. & Pathak, S. The protective role of Wisdom in Suicidal behavior: evidence from Indian adults. BMC Psychol 13, 1191 (2025).

35. Van Patten, R. et al. Assessment of 3-dimensional wisdom in schizophrenia: Associations with neuropsychological functions and physical and mental health. Schizophr Res 208, 360–369 (2019).

36. Bause, E. & Burbach, M. Purification and enzymatic properties of endo-alpha 1, 2-mannosidase from pig liver involved in oligosaccharide processing. Biol Chem 377, 639–646 (1996).

37. Jensen, K. P. et al. The α-endomannosidase gene (MANEA) is associated with panic disorder and social anxiety disorder. Transl Psychiatry 4, e353 (2014).

38. Farrer, L. A. et al. Association of variants in MANEA with cocaine-related behaviors. Arch Gen Psychiatry 66, 267–274 (2009).

39. Boison, D. Adenosine kinase: exploitation for therapeutic gain. Pharmacol Rev 65, 906–943 (2013).

40. Huang, J. et al. Genomics and phenomics of body mass index reveals a complex disease network. Nat Commun 13, 7973 (2022).

41. Pulit, S. L. et al. Meta-analysis of genome-wide association studies for body fat distribution in 694 649 individuals of European ancestry. Hum Mol Genet 28, 166–174 (2019).

42. Sidorenko, J. et al. Genetic architecture reconciles linkage and association studies of complex traits. Nat Genet 56, 2352–2360 (2024).

43. Koskeridis, F. et al. Pleiotropic genetic architecture and novel loci for C-reactive protein levels. Nat Commun 13, 6939 (2022).

44. Grgic, O. et al. Novel Genetic Determinants of Dental Maturation in Children. J Dent Res 102, 349–356 (2023).

45. Geller, F. et al. Genome-wide association study identifies four loci associated with eruption of permanent teeth. PLoS Genet 7, e1002275 (2011).

46. Fatemifar, G. et al. Genome-wide association study of primary tooth eruption identifies pleiotropic loci associated with height and craniofacial distances. Hum Mol Genet 22, 3807–3817 (2013).

47. Willett, J. D. S. et al. Identification of 16 novel Alzheimer’s disease loci using multi-ancestry meta-analyses. Alzheimers Dement 21, e14592 (2025).

48. Ardelt, M. Disentangling the Relations Between Wisdom and Different Types of Well-Being in Old Age: Findings from a Short-Term Longitudinal Study. J Happiness Stud 17, 1963–1984 (2016).

49. Karlsson Linnér, R., et al. Genome-wide association analyses of risk tolerance and risky behaviors in over 1 million individuals identify hundreds of loci and shared genetic influences. Nat Genet 51, 245–257 (2019).

50. Wright, A. J. & Jackson, J. J. Do changes in personality predict life outcomes? J Pers Soc Psychol 125, 1495–1518 (2023).

51. Human, L. J. et al. Is change bad? Personality change is associated with poorer psychological health and greater metabolic syndrome in midlife. J Pers 81, 249–260 (2013).

52. Mroczek, D. K. & Spiro, A., III. Personality change influences mortality in older men. Psychol Sci 18, 371–376 (2007).

53. Baselmans, B. M. L. et al. Multivariate genome-wide analyses of the well-being spectrum. Nat Genet 51, 445–451 (2019).

54. Gupta, P. et al. A genome-wide investigation into the underlying genetic architecture of personality traits and overlap with psychopathology. Nat Hum Behav 8, 2235–2249 (2024).

55. Nagel, M. et al. Meta-analysis of genome-wide association studies for neuroticism in 449,484 individuals identifies novel genetic loci and pathways. Nat Genet 50, 920–927 (2018).

56. Lo, M.-T. et al. Genome-wide analyses for personality traits identify six genomic loci and show correlations with psychiatric disorders. Nat Genet 49, 152–156 (2017).

57. Delhom, I., Satorres, E. & Meléndez, J. C. Emotional intelligence intervention in older adults to improve adaptation and reduce negative mood. Int Psychogeriatr 34, 79–89 (2022).

58. Schonert-Reichl, K. A. et al. Enhancing cognitive and social-emotional development through a simple-to-administer mindfulness-based school program for elementary school children: a randomized controlled trial. Dev Psychol 51, 52–66 (2015).

59. Jennings, P. A. et al. Impacts of the CARE for Teachers program on teachers’ social and emotional competence and classroom interactions. Journal of Educational Psychology 109, 1010–1028 (2017).

60. Jazaieri, H. et al. Enhancing Compassion: A Randomized Controlled Trial of a Compassion Cultivation Training Program. J Happiness Stud 14, 1113–1126 (2013).

61. Haehner, P., Wright, A. J. & Bleidorn, W. A systematic review of volitional personality change research. Commun Psychol 2, 115 (2024).

62. Sternberg, R. J. Implicit theories of intelligence, creativity, and wisdom. Journal of Personality and Social Psychology 49, 607–627 (1985).

63. Staudinger, U. M. & Glück, J. Psychological wisdom research: commonalities and differences in a growing field. Annu Rev Psychol 62, 215–241 (2011).

64. Staudinger, U. M., Lopez, D. F. & Baltes, P. B. The Psychometric Location of Wisdom-Related Performance: Intelligence, Personality, and more? Pers Soc Psychol Bull 23, 1200–1214 (1997).

65. Sirin, S. R. Socioeconomic Status and Academic Achievement: A Meta-Analytic Review of Research. Review of Educational Research 75, 417–453 (2005).

66. Tucker-Drob, E. M., Briley, D. A., Engelhardt, L. E., Mann, F. D. & Harden, K. P. Genetically-mediated associations between measures of childhood character and academic achievement. J Pers Soc Psychol 111, 790–815 (2016).

67. Poropat, A. E. A meta-analysis of the five-factor model of personality and academic performance. Psychol Bull 135, 322–338 (2009).

68. Demange, P. A. et al. Investigating the genetic architecture of noncognitive skills using GWAS-by-subtraction. Nat Genet 53, 35–44 (2021).

69. Watson, C. J. et al. Splitting schizophrenia: divergent cognitive and educational outcomes revealed by genomic structural equation modelling. Mol Psychiatry 31, 3098–3107 (2026).

70. Mallard, T. T. et al. Item-Level Genome-Wide Association Study of the Alcohol Use Disorders Identification Test in Three Population-Based Cohorts. Am J Psychiatry 179, 58–70 (2022).

71. Takahashi, M. & Bordia, P. The Concept of Wisdom: A Cross-cultural Comparison. Int J Psychol 35, 1–9 (2000).

72. Sanchez-Roige, S. et al. Genome-wide association study of delay discounting in 23,217 adult research participants of European ancestry. Nat Neurosci 21, 16–18 (2018).

73. Sanchez-Roige, S. et al. Genome-wide association study of problematic opioid prescription use in 132,113 23andMe research participants of European ancestry. Mol Psychiatry 26, 6209–6217 (2021).

74. Sanchez-Roige, S. et al. CADM2 is implicated in impulsive personality and numerous other traits by genome- and phenome-wide association studies in humans and mice. Transl Psychiatry 13, 167 (2023).

75. Thorpe, H. H. A. et al. Genome-wide association study of delay discounting identifies 11 loci and reveals transdiagnostic associations across mental and physical health. Mol Psychiatry 31, 2081–2093 (2026).

76. Thorpe, H. H. A. et al. Genome-wide association studies of lifetime and frequency of cannabis use in 131,895 individuals. Mol Psychiatry 31, 1061–1073 (2026).

77. Sollis, E. et al. The NHGRI-EBI GWAS Catalog: knowledgebase and deposition resource. Nucleic Acids Res 51, D977–D985 (2023).

78. Wellcome Trust Case Control Consortium et al. Bayesian refinement of association signals for 14 loci in 3 common diseases. Nat Genet 44, 1294–1301 (2012).

79. Winkler, T. W. et al. Quality control and conduct of genome-wide association meta-analyses. Nat Protoc 9, 1192–1212 (2014).

80. Willer, C. J., Li, Y. & Abecasis, G. R. METAL: fast and efficient meta-analysis of genomewide association scans. Bioinformatics 26, 2190–2191 (2010).

81. Chang, C. C. et al. Second-generation PLINK: rising to the challenge of larger and richer datasets. Gigascience 4, 7 (2015).

82. 1000 Genomes Project Consortium et al. A global reference for human genetic variation. Nature 526, 68–74 (2015).

83. Yang, J., Lee, S. H., Goddard, M. E. & Visscher, P. M. GCTA: a tool for genome-wide complex trait analysis. Am J Hum Genet 88, 76–82 (2011).

84. de Leeuw, C. A., Mooij, J. M., Heskes, T. & Posthuma, D. MAGMA: generalized gene-set analysis of GWAS data. PLoS Comput Biol 11, e1004219 (2015).

85. Watanabe, K., Taskesen, E., van Bochoven, A. & Posthuma, D. Functional mapping and annotation of genetic associations with FUMA. Nat Commun 8, 1826 (2017).

86. Barbeira, A. N. et al. Exploring the phenotypic consequences of tissue specific gene expression variation inferred from GWAS summary statistics. Nat Commun 9, 1825 (2018).

87. Bulik-Sullivan, B. et al. An atlas of genetic correlations across human diseases and traits. Nat Genet 47, 1236–1241 (2015).

88. Bulik-Sullivan, B. K. et al. LD Score regression distinguishes confounding from polygenicity in genome-wide association studies. Nat Genet 47, 291–295 (2015).

89. All of Us Research Program Investigators et al. The ‘All of Us’ Research Program. N Engl J Med 381, 668–676 (2019).

90. All of Us Research Program Genomics Investigators. Genomic data in the All of Us Research Program. Nature 627, 340–346 (2024).

91. Ramirez, A. H. et al. The All of Us Research Program: Data quality, utility, and diversity. Patterns (N Y) 3, 100570 (2022).

92. Martin, A. R. et al. Clinical use of current polygenic risk scores may exacerbate health disparities. Nat Genet 51, 584–591 (2019).

93. Ge, T., Chen, C.-Y., Ni, Y., Feng, Y.-C. A. & Smoller, J. W. Polygenic prediction via Bayesian regression and continuous shrinkage priors. Nat Commun 10, 1776 (2019).

