## Supplementary Information for "The genetic architecture of wisdom and its relationship with health, personality and cognition"

### Supplementary Methods

*Analysis of local ancestry*

We performed total wisdom score GWAS on research-consented participants with the 23andMe Research Institute. Ancestry falls along a spectrum[1,2]. Participants were classified as being of European-like ($N_{EUR}$=131,870), Latin American-like ($N_{LAT}$=16,812), and African-like ($N_{AFR}$=4,702) genetic ancestry. Individuals were clustered by genetic similarity to a reference panel using local ancestry analysis[3]. The 23andMe algorithm first partitions phased genomic data into short windows of about 300 SNPs. Within each window, a support vector machine classifies individual haplotypes into one of 31 reference populations (https://www.23andme.com/ancestry-composition-guide/). The support vector machine classifications are fed into a hidden Markov model that accounts for switch errors and incorrect assignments, and gives probabilities for each reference population in each window. The reference population data is derived from public datasets (the Human Genome Diversity Project, HapMap, and 1000 Genomes), as well as 23andMe research participants who have reported having four grandparents from the same country.

Ancestries are defined as follows:

| **Ancestry** | **Classification Criteria** |
|---|---|
| European | European + Middle Eastern > 0.97, European > 0.90 |
| African American + Latin American | European + African + East Asian + Native American + Middle Eastern > 0.90, African + Native American > 0.01 |

**Genome-wide association and secondary analyses**

DNA extraction and genotyping were performed from saliva samples by clinical laboratories CLIA-certified and CAP-accredited by the Laboratory Corporation of America. The 23andMe Research Institute conducted quality control, imputation, and univariate genome-wide analyses as previously described[4,5]. Participants were genotyped on one of five Illumina genotyping platforms, containing between 550,000 to 950,000 variants, with 1,507,585 genotyped variants entering the variant-level QC summaries. Samples were genotyped on one of five genotyping platforms. The V1 and V2 platforms were variants of the Illumina HumanHap550 + BeadChip, including about 25,000 custom SNPs selected by 23andMe, with a total of ~560,000 SNPs. The V3 platform was based on the Illumina OmniExpress + BeadChip, with custom content to improve the overlap with our V2 array, with a total of ~950,000 SNPs. The V4 platform is a fully custom array, including a lower redundancy subset of V2 and V3 SNPs with additional coverage of lower-frequency coding variation, and ~570,000 SNPs. The v5 platform, in current use, is an Illumina Infinium Global Screening Array (~640,000 variants) supplemented with ~50,000 variants of custom content. Samples that failed to reach 98.5% call rate were first re-analyzed, then excluded from the study. Variants were imputed based on an imputation panel combining 1000 Genomes Phase 3, UK10K and the Human Reference Consortium (**HRC**). About 64.4 million variants were then imputed against the HRC panel, augmented by a single unified imputation reference panel combining the May 2015 release of the 1000 Genomes Phase 3 haplotypes with the UK10K imputation reference panel for variants not present in the HRC. Imputed variants with low imputation quality ($r^2 < 0.50$ averaged across batches or a minimum $r^2 < 0.30$), or with evidence of batch effects ($p <$ 1.00E-50) were removed. We filtered out variants with minor allele frequency (**MAF**) < 0.1%, which are extremely sensitive to quantitative trait over-dispersion. Principal components were computed using ~65,000 high-quality genotyped variants present in all five genotyping platforms.

A maximal set of unrelated individuals was chosen for the analysis using a segmental identity-by-descent (**IBD**) estimation algorithm[6]. Individuals were defined as related if they shared more than 700 cM IBD, including regions where the two individuals shared either one or both genomic segments IBD. This level of relatedness (~20% of the genome) corresponds to approximately the minimal expected sharing between first cousins in an outbred population. The 23andMe GWAS pipeline performs linear regression and assumes an additive model for allelic effects[7–11]. Age, sex, the top five principal genotype components in the European-like cohort, nine in Latin American-like cohort, and six in the African-like cohort, and indicator variables for genotype platforms were applied as covariates. *P*-values were corrected for genomic inflation. We imputed participant genotype data against the September 2013 release of 1000 Genomes phase 3 reference haplotypes. Phasing and imputation were performed for each genotyping platform separately. We phased using an internally developed phasing tool, Finch, which implements the Beagle haplotype graph-based phasing algorithm, modified to separate the haplotype graph construction and phasing steps. In preparation for imputation, we split phased chromosomes into segments of no more than 10,000 genotyped SNPs, with overlaps of 200 SNPs. We excluded SNPs of low genotyping quality, including those that failed a Mendelian transmission test in trios ($p < 1.00\text{E-}20$) or with large allele frequency discrepancies compared to reference genome data, failed Hardy-Weinberg test ($p < 1.00\text{E-}20$), failed batch effects test (ANOVA $p < 1.00\text{E-}20$), or had a call rate < 90%. Frequency discrepancies were identified by computing a 2 x 2 table of allele counts for European 1000 Genomes samples and 2000 randomly sampled 23andMe research participants with European-like ancestries, and identifying SNPs with a $\chi^2$ $p < 1.00\text{E-}15$. We imputed each phased segment against all-ethnicity 1000 Genomes haplotypes (excluding monomorphic and singleton sites) using Minimac2[12], using 5 rounds and 200 states for parameter estimation.

For the X chromosome, we built separate haplotype graphs for the non-pseudoautosomal region and each pseudoautosomal region. These regions were phased separately. We then imputed males and females together using Minimac2, as with the autosomes, treating males as homozygous pseudo-diploids for the non-pseudoautosomal region.

For tests using imputed data, we use the imputed dosages rather than best-guess genotypes. We imputed HLA allele dosages from SNP genotype data using HIBAG. We imputed alleles for HLA-A, B, C, DPB1, DQA1, DQB1, and DRB1 loci at four-digit resolution. To test associations between HLA allele dosages and phenotypes, we performed linear regression using the same set of covariates used in the SNP based GWAS. We performed separate association tests for each imputed allele. HLA analysis did not identify any significant signal.

**Hi-C coupled Multi-marker Analysis of GenoMic Annotation**

We incorporated total wisdom score GWAS data with chromatin interaction profiles from the human brain using Hi-C coupled MAGMA (**H-MAGMA**)[13]. H-MAGMA assigns non-coding SNPs to genes based on chromatin interactions from fetal brain, adult brain, midbrain neurons, cortical neurons, iPSC-derived neurons, and iPSC-derived astrocytes datasets (https://github.com/thewonlab/H-MAGMA). Exonic and promoter SNPs were assigned to genes based on physical position[13]. We applied correction based on the total number of gene-tissue pairs tested ($N_{genes}$ per tissue = 53,046 to 53,320; Bonferroni $p$ < 9.38E-07 to 9.43E-07).

**Gene-set and Tissue Enrichment Analysis**

Gene-set and tissue enrichment analyses analysis was conducted using MAGMA through the web-based platform Functional Mapping and Annotation of Genome-Wide Association Studies (FUMA v1.8.3)[14]. SNPs were annotated to protein-coding genes with Ensembl build v110. The default settings (SNP-wise mean model, MAF = 0.01%, 0kb gene window, sex as a covariate,

Positional mapping parameters: 10kb distance) were used. LD was estimated using the 1000 Genomes European reference sample. Gene-set analysis was conducted on 17,011 gene-sets with Gene Ontology terms curated from the Molecular Signatures Database (MsigDB v7.0). Significance was determined at a Bonferroni threshold based on the total number of gene-sets tested ($p$ < 2.94E-06). Tissue-specific gene expression profiles for 54 tissues were assessed. Average gene expression in each tissue used as a covariate. Using GTEx v8 RNA-seq data, gene expression values were $\log_2$ transformed from the average Reads Per Kilobase Million (max value = 50) per tissue. Significance was determined following Bonferroni correction for each tissue comparison ($p$ < 9.26E−04).

**Cell-Type Enrichment Analysis**

We tested associations between cell types associated with total wisdom score according to Duncan et al. 2025[15]. Briefly, MAGMA results identified with GWAS data were integrated with single nuclei RNA sequencing data[16] to characterize brain cell types that may be most relevant to wisdom. The cluster-by-gene matrix and specificity scores to quantify gene expression in 461 cell types were derived from the provided specificity score matrix (https://github.com/Integrative-Mental-Health-Lab/linking_cell_types_to_brain_phenotypes/). MAGMA was used to map wisdom-associated SNPs to genes located within 35 kb upstream to 10 kb downstream of the SNP. Linear regression analysis in MAGMA was used to test relationships between specificity scores and total wisdom score association for genes, with each cell type analyzed independently. Analysis was conducted with the SNP-wise mean model adjusted for LD derived from the 1000G phase 3 European ancestry panel.

**Phenome-Wide Association Study**

Taking a hypothesis-free approach to assess if wisdom genetics were associated with various health outcomes, we conducted a phenome-wide association study (**PheWAS**). Total wisdom score polygenic scores (**PGS**) calculated in those of European-like genetic ancestry were associated with diagnoses spanning physical and mental health available from electronic health records (**EHR**) in the All of Us (**AoU**) cohort (version 8). See main text for PGS calculation. For PheWAS, we fitted a logistic regression model to each case/control disease phenotype ("phecode") to estimate the log odds of each diagnosis given total wisdom score PGS, while adjusting for sex at birth, age at last EHR event, and the first 5 PCs using the Python package PheTK (v0.2.12)[17]. At least two **ICD**-code occurrences mapping to a PheWAS disease category (Phecode Map version 1.2; https://phewascatalog.org/phecodes) were required for case classification, and a minimum of 100 cases and 100 controls were required for phecode inclusion. EHR duration was generated but omitted from the fitted covariates after a column-detection warning. The covariate-complete cohort included 155,547 participants. Of 1,813 phecodes present, 210 failed the minimum case/control filter, 1,603 tests were attempted, and 1,600 converged. We calculated the 5% FDR for all associations performed ($N$ = 1,600; **Supplementary Table 25**).

**Factor Analysis**

To examine the loadings of individual SD-WISE-7 items on overall wisdom, we fitted confirmatory factor analysis (**CFA**) models with a unidimensional factor structure using the semopy package (version 2.3.11) in Python. Parameters were estimated using maximum likelihood (**MLW**), treating items as continuous. We compared a one-factor model with a modified model allowing a post hoc residual covariance between Self-Reflection and Acceptance of Divergent Perspectives. Both models were also refitted in two random halves of the sample as a stability check. Comparative fit index (**CFI**) and root mean square error of approximation (**RMSEA**) statistics were used to evaluate model fit. A CFI $\geq 0.90$ and a SRMR $< 0.10$ are generally considered acceptable. CFI above 0.95 and RMSEA values below 0.06 are considered excellent; above 0.90 and below 0.08, respectively, are considered adequate[18].

### **Supplementary Figures**

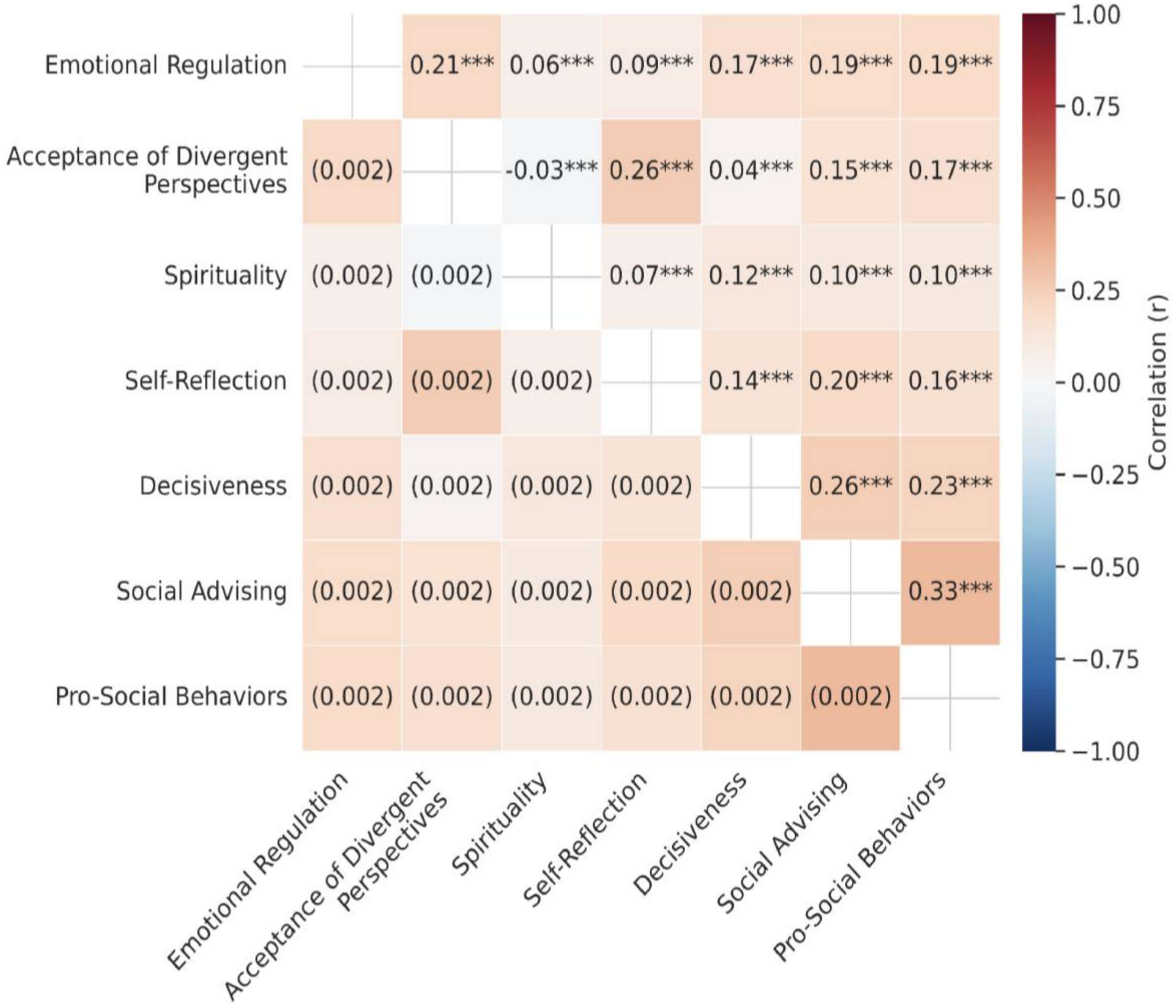


***Supplementary Figure 1.*** **Inter-item phenotypic correlations.** Unadjusted inter-item phenotypic correlations across SD-WISE-7 items ($N$ = 161,775). The upper triangle shows Pearson $r$ and significance (***uncorrected $p < 0.001$); the lower triangle shows the standard error. See **Supplementary Table 3** for full dataset.

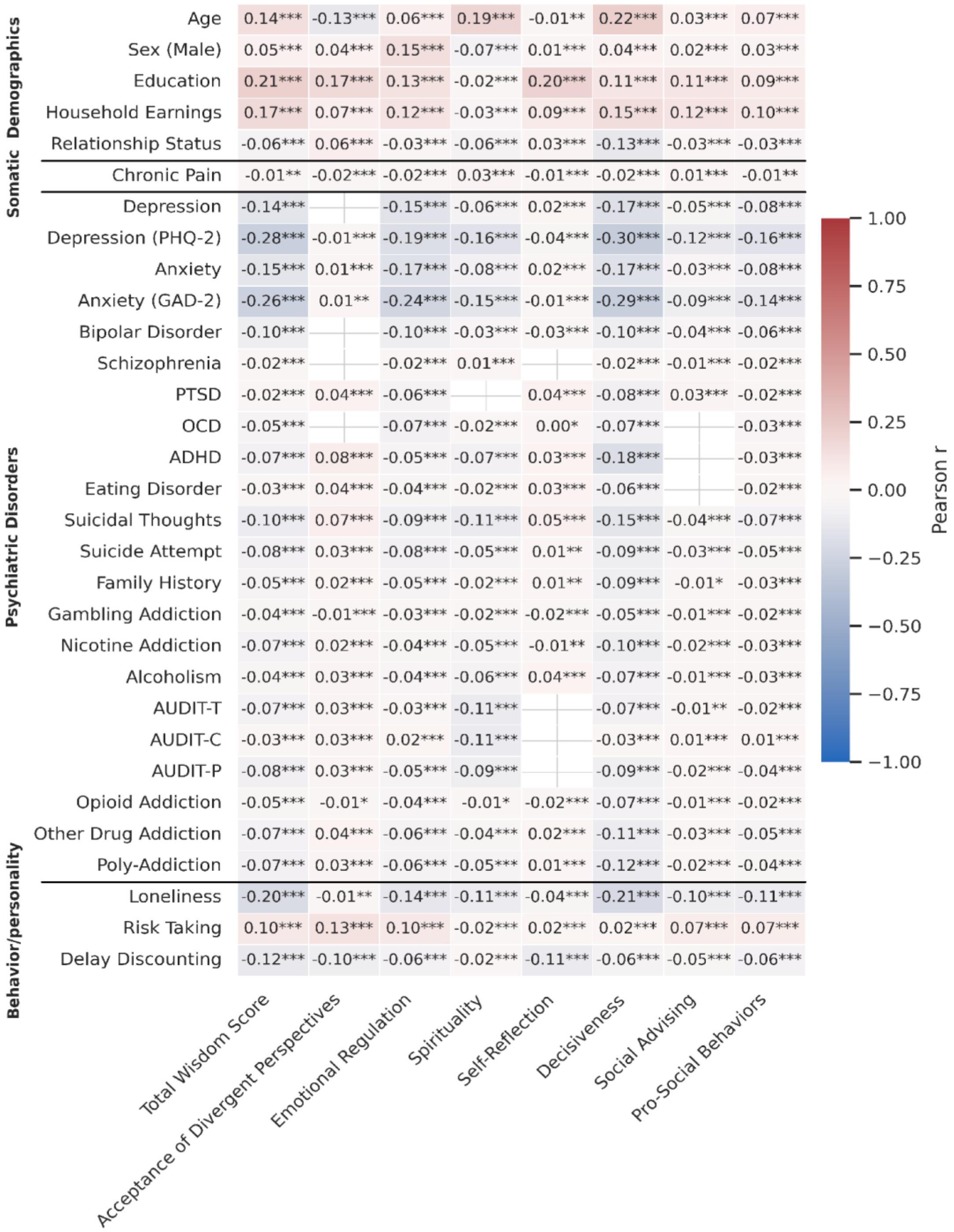


***Supplementary Figure 2.*** **Cross-trait item phenotypic correlations.** Unadjusted Pearson correlations of the total wisdom score and the seven individual wisdom items with 31 selected traits. For sex correlations, male = 1 and female = 0. See **Supplementary Table 4**.

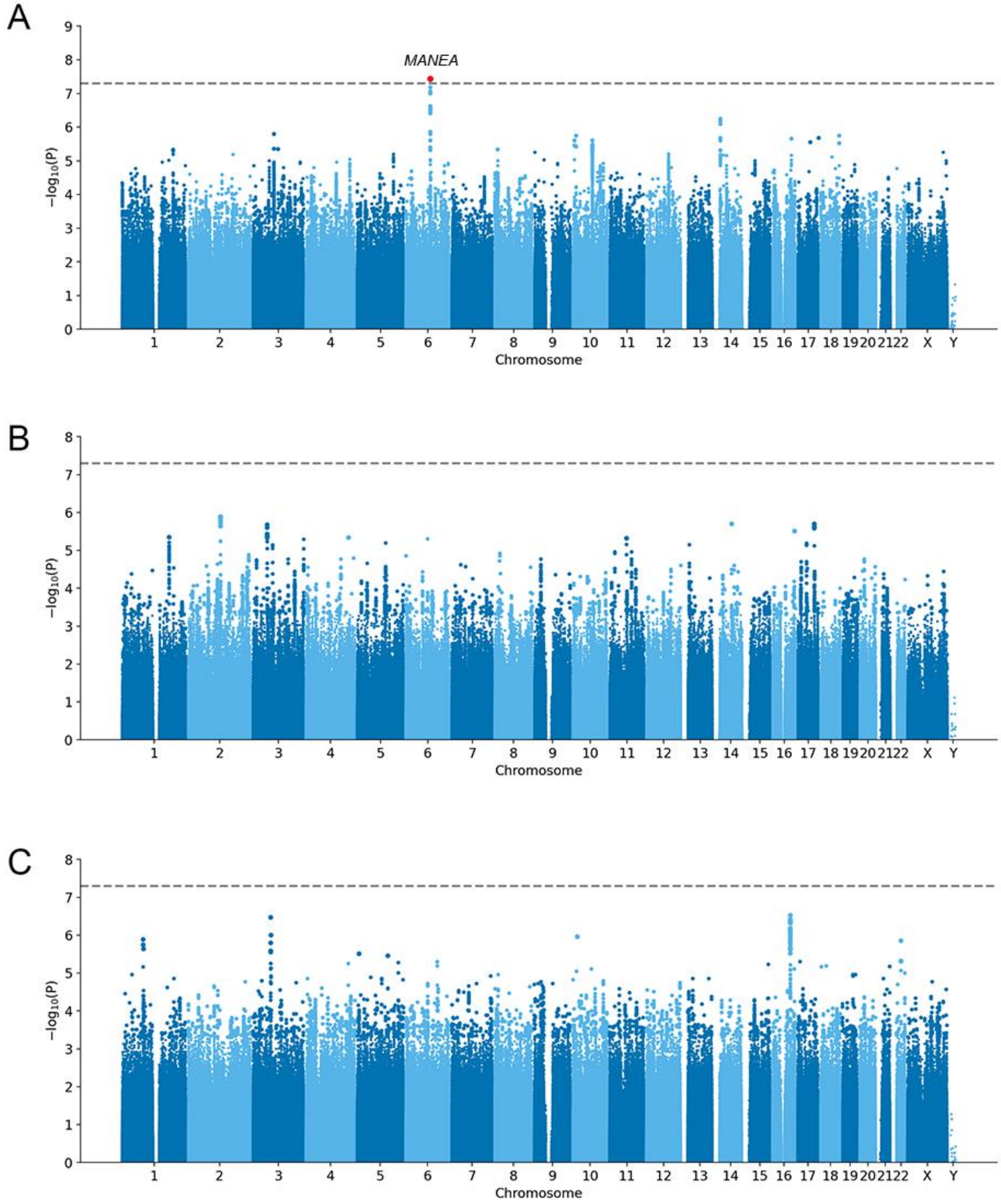


***Supplementary Figure 3.*** **Individual ancestry GWAS**. GWAS of total wisdom scores in **A)** European-, **B)** Latin American-, and **C)** African-like genetic ancestry participants, after downstream common-variant QC (MAF ≥ 1%). The red marker highlights the EUR lead variant, and the dashed line marks $p$ = 5.00E-08. The locus is labelled with the nearest gene.

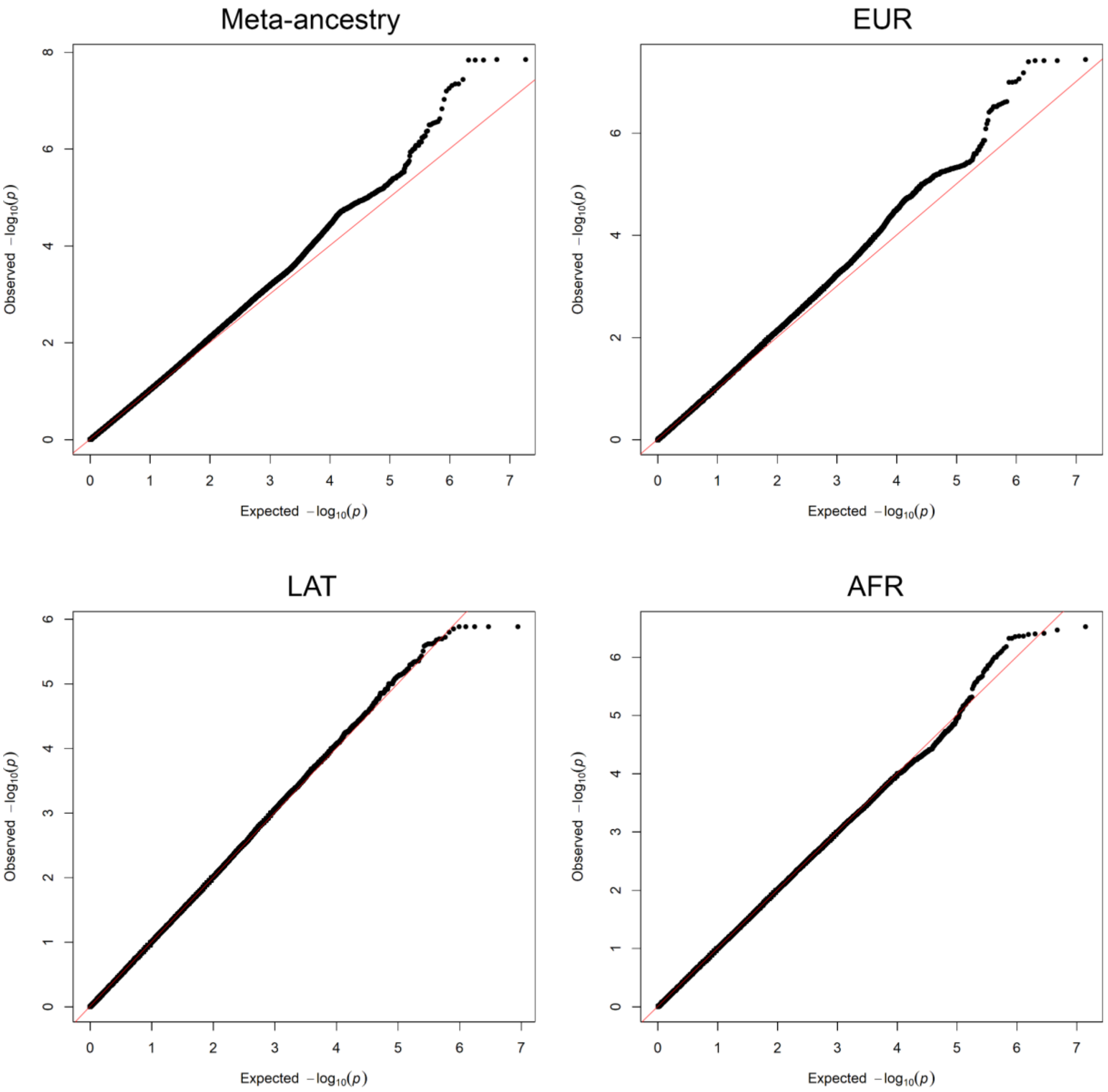


***Supplementary Figure 4*****. Q-Q plots.** Observed versus expected GWAS p values for the multi-ancestry meta-analysis, European-like ancestry (**EUR**), Latin American-like ancestry (**LAT**), and African American-like ancestry (**AFR**).

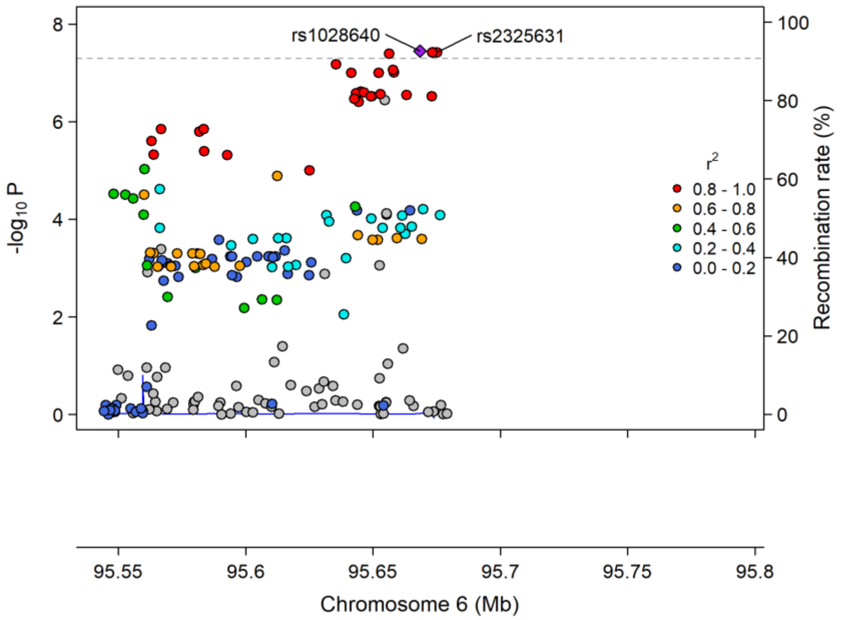


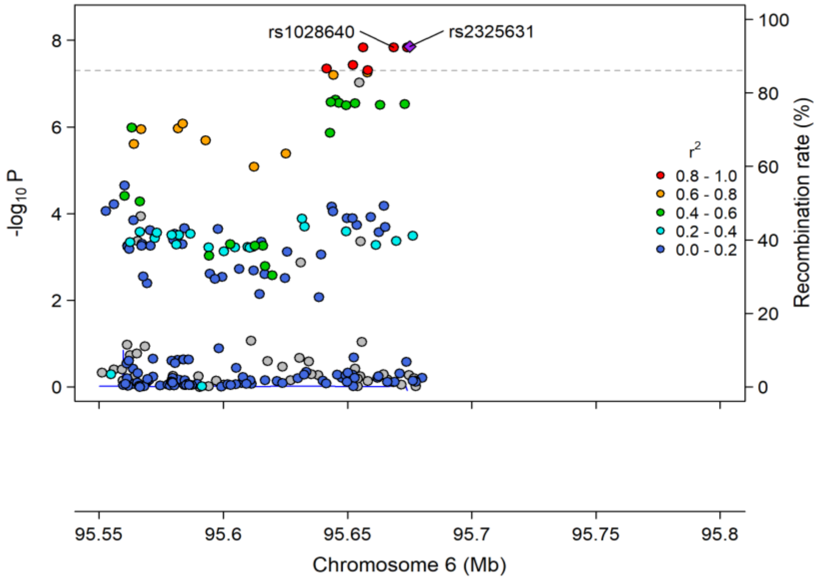


***Supplementary Figure 5.*** **Locus zoom plots.** European-like ancestry locus plot (top) and meta-analysis (bottom) locus zoom plot of the chromosome 6 locus. The EUR lead is rs1028640 at chromosome 6:95,668,509 (GRCh37; GRCh38 position 94,958,792), at the same locus as the meta-analysis signal.

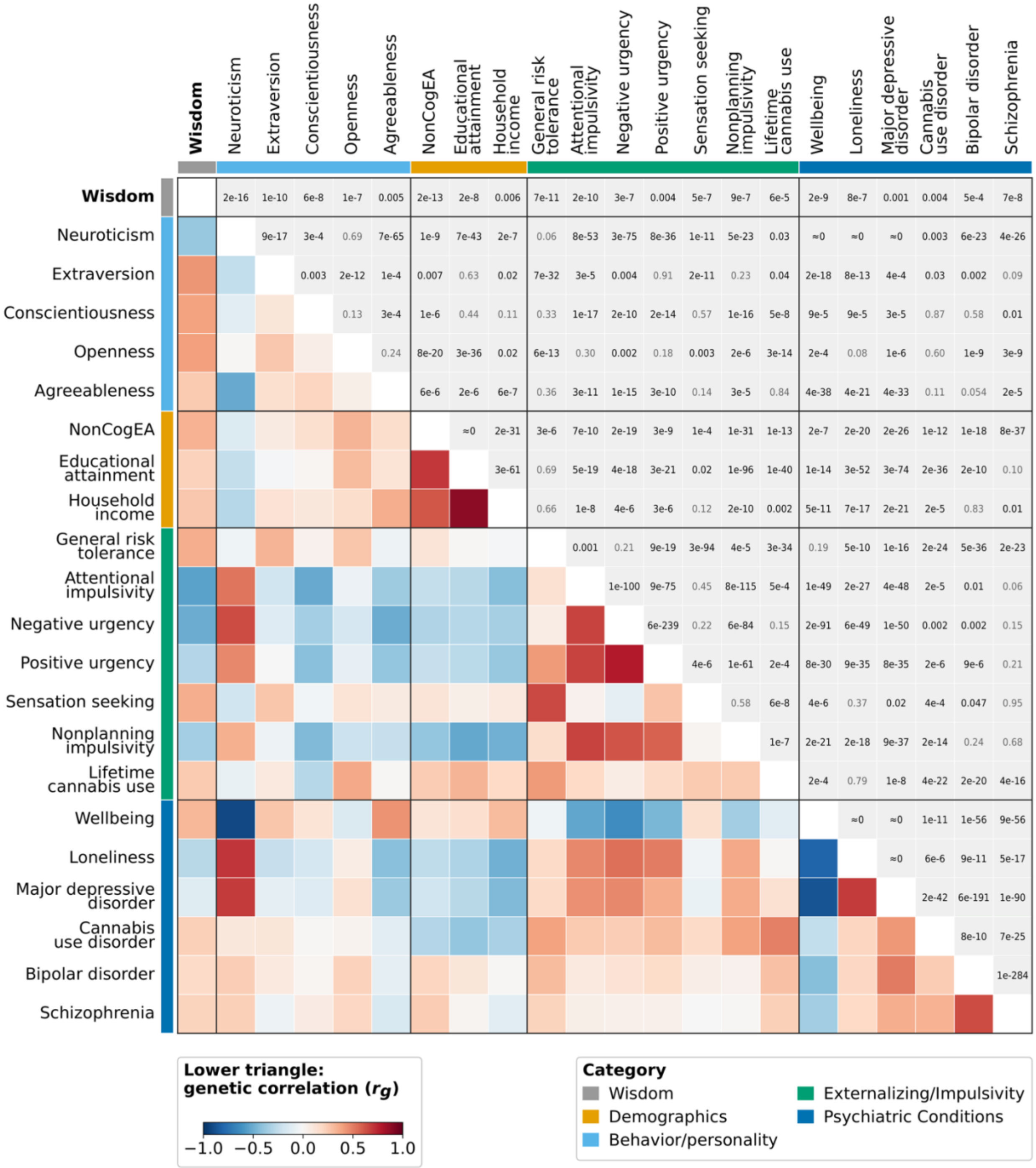


***Supplementary Figure 6*. Genetic-correlation matrix.** Pairwise genetic correlations ($r_g$) among total wisdom scores and 21 significant traits after FDR correction. The lower triangle shows $r_g$ and the upper triangle shows uncorrected pairwise $p$ values. See **Supplementary Table 16** for additional details.

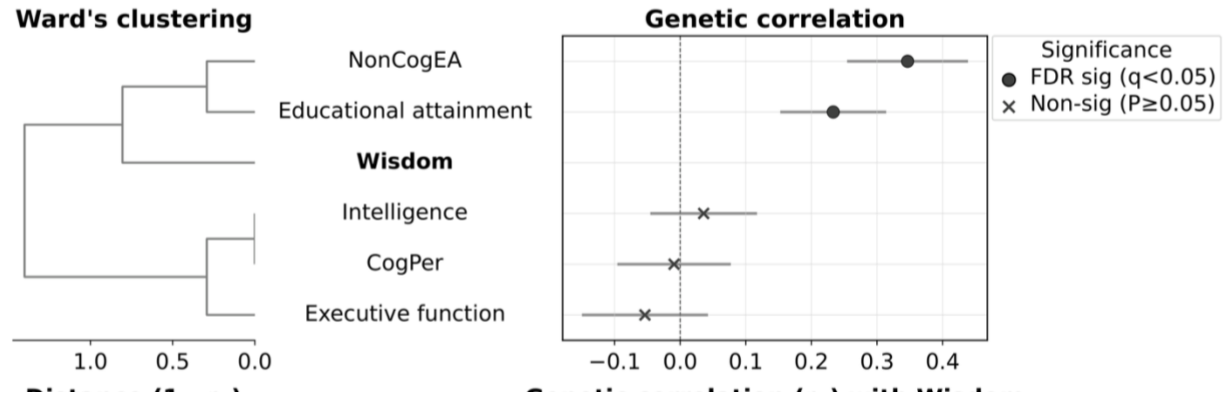


***Supplementary Figure 7.*** **Cognitive-trait genetic correlations.** Genetic correlations of wisdom with cognitive traits, with hierarchical clustering.

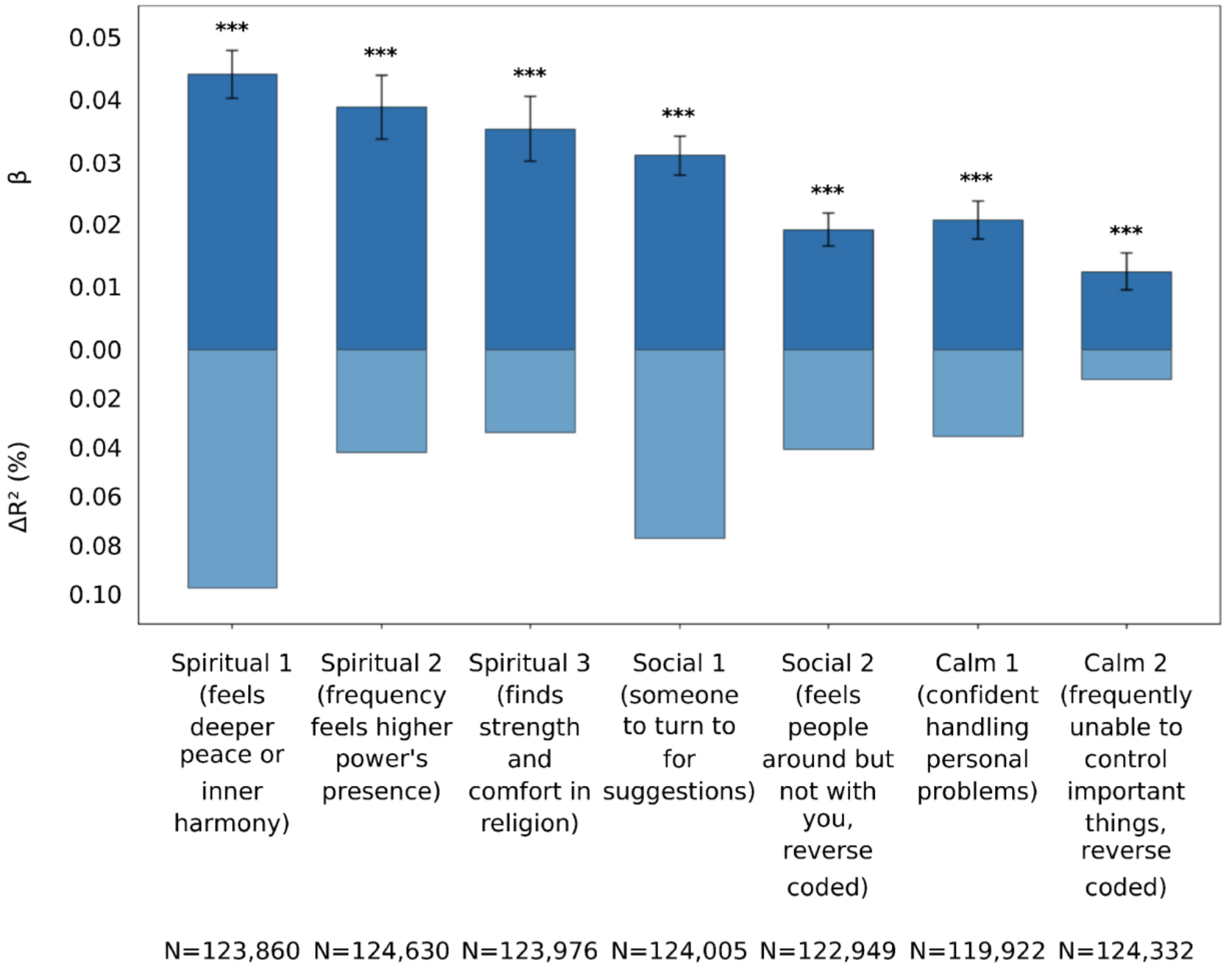


***Supplementary Figure 8.*** **AoU SD-WISE analogue PGS.** Associations between total wisdom score PGS and seven AoU survey items serving as proxies of the Spirituality, Pro-Social Behaviors, and Emotional Regulation items of SD-WISE-7 ($N_{EUR} \leq$ 124,630). Models include age, sex, PC1–10 and ZSDI. Top bars show the effect (**β**), expressed in outcome units per standard deviation of wisdom PGS, ± standard error. Bottom bars show variance explained by PGS (**$\Delta R^2$**). Asterisks denote FDR significance across seven items (***$q$ < 0.001). See **Supplementary Table 18**.

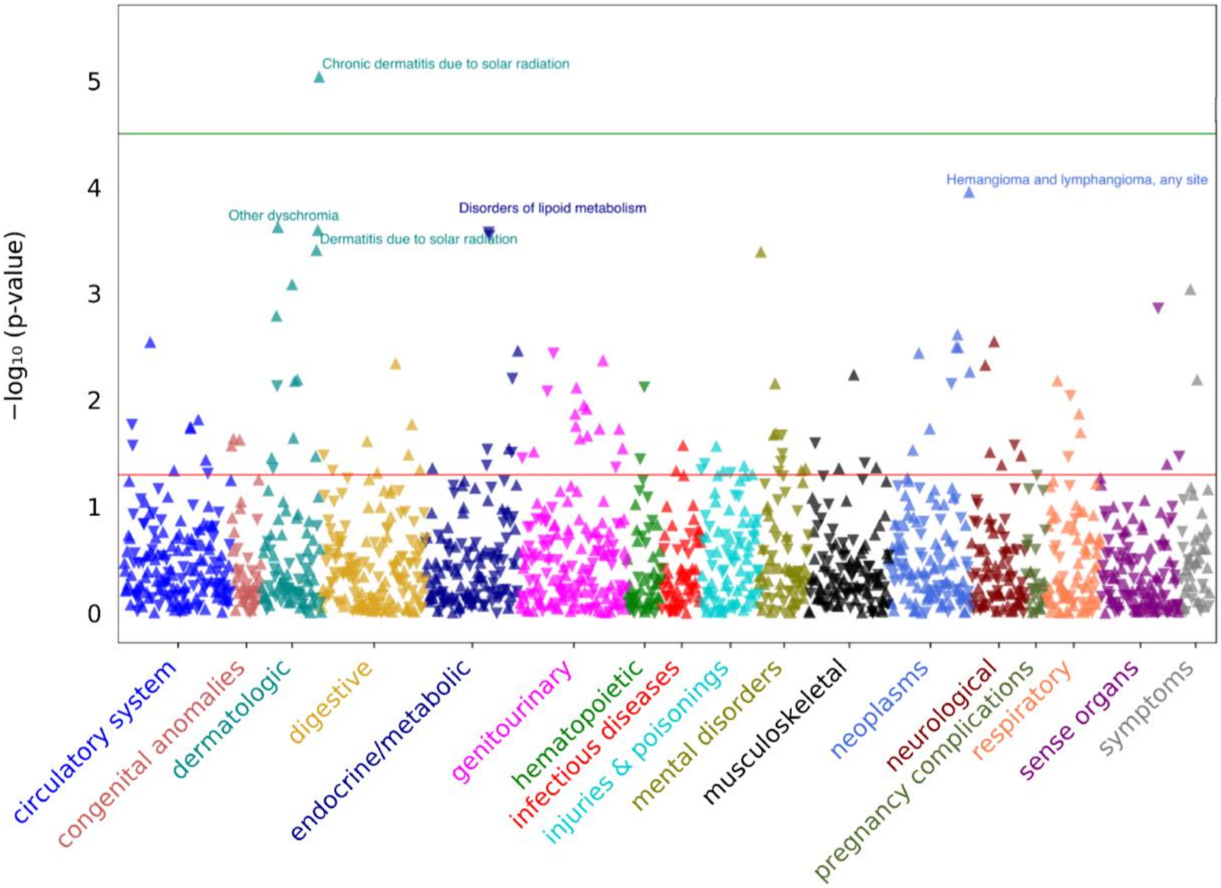


***Supplementary Figure 9.*** **AoU PheWAS.** Points show 1,600 converged diagnostic tests. The red line marks nominal significance ($p$ = 0.05) and green line indicates Bonferroni significance ($p$ = 3.13E-05). One association survived correction (**Supplementary Table 25**).

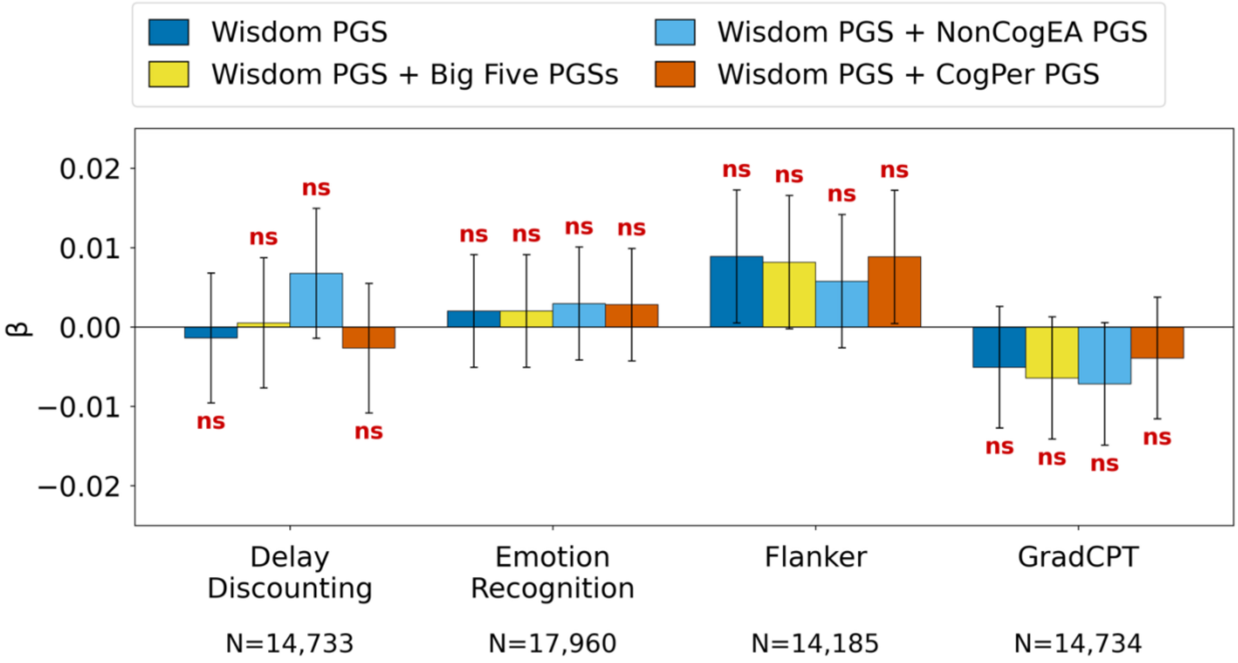


***Supplementary Figure 10*. Wisdom PGS associations with cognitive task performance.** Associations between wisdom PGS and performance on four cognitive tasks in All of Us participants ($N_{EUR} \leq$ 17,960): Delay Discounting, Emotion Recognition/Guess the Emotion, Flanker, and the Gradual Onset Continuous Performance Task (**GradCPT**). Bars show total wisdom score PGS standardized effect (β) ± standard error. Models include the main model (age, sex, PC1-10, ZSDI, wisdom PGS); the main model plus all Big Five PGSs; the main model plus NonCogEA PGS; and the main model plus cognitive performance (**CogPer**) PGS. ns = non-significant. See **Supplementary Tables 26-31** for model outputs.

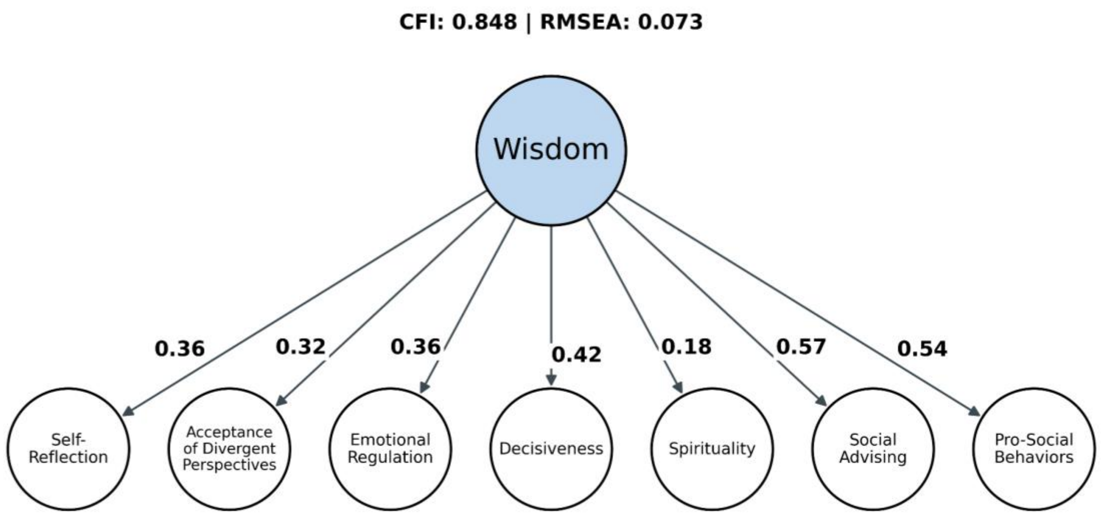


***Supplementary Figure 11.*** **Common factor model**. SD-WISE-7 data from 161,775 complete cases fitted to a unidimensional factor structure.

### References


1. Bryc, K., Durand, E. Y., Macpherson, J. M., Reich, D. & Mountain, J. L. The genetic ancestry of African Americans, Latinos, and European Americans across the United States. *Am J Hum Genet* **96**, 37–53 (2015).
2. National Academies of Sciences, Engineering, and Medicine. *Using Population Descriptors in Genetics and Genomics Research: A New Framework for an Evolving Field*. (The National Academies Press, Washington, DC, 2023). doi:10.17226/26902.
3. Durand, E. Y., Do, C. B., Mountain, J. L. & Macpherson, J. M. Ancestry Composition: A Novel, Efficient Pipeline for Ancestry Deconvolution. Preprint at https://doi.org/10.1101/010512 (2014).
4. Eriksson, N. *et al.* Web-based, participant-driven studies yield novel genetic associations for common traits. *PLoS Genet* **6**, e1000993 (2010).
5. Hyde, C. L. *et al.* Identification of 15 genetic loci associated with risk of major depression in individuals of European descent. *Nat Genet* **48**, 1031–1036 (2016).
6. Henn, B. M. *et al.* Cryptic distant relatives are common in both isolated and cosmopolitan genetic samples. *PLoS One* **7**, e34267 (2012).
7. Sanchez-Roige, S. *et al.* Genome-wide association study of delay discounting in 23,217 adult research participants of European ancestry. *Nat Neurosci* **21**, 16–18 (2018).
8. Sanchez-Roige, S. *et al.* Genome-wide association study of problematic opioid prescription use in 132,113 23andMe research participants of European ancestry. *Mol Psychiatry* **26**, 6209–6217 (2021).
9. Sanchez-Roige, S. *et al.* CADM2 is implicated in impulsive personality and numerous other traits by genome- and phenome-wide association studies in humans and mice. *Transl Psychiatry* **13**, 167 (2023).

10. Thorpe, H. H. A. *et al.* Genome-wide association study of delay discounting identifies 11 loci and reveals transdiagnostic associations across mental and physical health. *Mol Psychiatry* **31**, 2081–2093 (2026).
11. Thorpe, H. H. A. *et al.* Genome-wide association studies of lifetime and frequency of cannabis use in 131,895 individuals. *Mol Psychiatry* **31**, 1061–1073 (2026).
12. Fuchsberger, C., Abecasis, G. R. & Hinds, D. A. minimac2: faster genotype imputation. *Bioinformatics* **31**, 782–784 (2015).
13. Sey, N. Y. A. *et al.* A computational tool (H-MAGMA) for improved prediction of brain-disorder risk genes by incorporating brain chromatin interaction profiles. *Nat Neurosci* **23**, 583–593 (2020).
14. Watanabe, K., Taskesen, E., van Bochoven, A. & Posthuma, D. Functional mapping and annotation of genetic associations with FUMA. *Nat Commun* **8**, 1826 (2017).
15. Duncan, L. E. *et al.* Mapping the cellular etiology of schizophrenia and complex brain phenotypes. *Nat Neurosci* **28**, 248–258 (2025).
16. Siletti, K. *et al.* Transcriptomic diversity of cell types across the adult human brain. *Science* **382**, eadd7046 (2023).
17. Tran, T. C. *et al.* PheWAS analysis on large-scale biobank data with PheTK. *Bioinformatics* **41**, btae719 (2025).
18. Hu, L. & Bentler, P. M. Cutoff criteria for fit indexes in covariance structure analysis: Conventional criteria versus new alternatives. *Structural Equation Modeling: A Multidisciplinary Journal* **6**, 1–55 (1999).
